# Unified Machine Learning Model for Early Detection of Mild Cognitive Impairment, Frailty, and Fall Risk Using Multimodal Motor-Function Data

**DOI:** 10.64898/2026.11.19.26368462

**Authors:** Sonia Akter, Trent M. Guess, Shraboni Sarker, Samuel A. Hockett, Andrew M. Kiselica, Jamie B. Hall, Praveen Rao

## Abstract

**Background:** Mild cognitive impairment (MCI), frailty, and fall risk often co-occur in older adults and share motor signs, yet they are screened and modeled separately. A unified model that predicts all 3 conditions from 1 sensor-based assessment could support earlier and more efficient screening, but such a model has rarely been developed or rigorously compared with separate models.

**Objective:** This study aimed to develop a unified machine learning model that predicts MCI, frailty, and fall risk together from sensor-based motor-function data, compare it with separate single-label models, explain its predictions, and test its robustness.

**Methods:** Community-dwelling older adults (n=79) completed a multimodal motor-function assessment with the Mizzou Point-of-Care Assessment System (MPASS), covering gait, sit-to-stand, static balance, step-down, and reaction time, with some tasks repeated under a dual-task condition. With demographic and family-history data, this yielded 59 features. Unified classifier-chain and single-label models were built with 5 algorithms across 6 modeling stages, evaluated with repeated nested cross-validation, and compared using the area under the receiver operating characteristic curve (AUC) and paired bootstrap tests. The final model was explained with Shapley additive explanations (SHAP), and its robustness and the sufficiency of the sample size were tested with chain-input, chain-order sensitivity, domain-ablation, learning-curve, and power analyses.

**Results:** With class weighting, the unified model had a higher mean AUC than the single-label models in 17 of 20 configurations and was never significantly worse, and it performed at least as well under nested model selection (0.615 vs 0.605; P=.58). The final extra trees model achieved a mean AUC of 0.645 (95% CI 0.569-0.722); discrimination was best for MCI (AUC 0.779, 95% CI 0.680-0.872) and lower for frailty (0.606) and fall risk (0.551). The model overestimated risk for all 3 outcomes, most for frailty. Most features (43/59, 73%), led by stride length, were shared across the 3 conditions, with consistent SHAP rankings in training and test participants (ρ=0.99). Predicted frailty was the most important input for fall risk, and the 45 sensor features alone performed as well as the full model. Learning curves suggested that fall-risk prediction would benefit from more participants.

**Conclusions:** A unified model predicted MCI, frailty, and fall risk together from one sensor-based assessment and performed as well as 3 separate models. It identified motor features shared across the conditions and used the link between frailty and fall risk. It performed best for MCI; with improvement for frailty and fall risk, recalibration, and external validation, such a model could support efficient screening of older adults for several conditions at once.

## Introduction

Population aging is increasing the number of older adults living with conditions that threaten their independence and quality of life and add to the healthcare burden [1,2]. Three of the most common are mild cognitive impairment (MCI), frailty, and fall risk. MCI is a decline in memory or thinking beyond normal aging in which people still manage daily life independently, and it often progresses to dementia [3]. Frailty is a loss of physiological reserve that increases vulnerability to disability, hospitalization, and death [4,5]. Falls are a leading cause of injury and loss of independence in older adults [6]. Early detection of these conditions gives clinicians time to intervene before decline becomes severe.

These conditions could also be detected together, because they are closely related: they often occur in the same person and share underlying motor impairments [7–9]. Slower gait is associated with cognitive impairment and a higher risk of dementia [10–12], reduced walking speed and muscle weakness are key components of frailty [4], frailty predicts future falls [13], and older adults with both frailty and cognitive impairment fall more often than those with either condition alone [8,9]. In routine care, however, each condition is assessed separately, with a cognitive screening test, a frailty questionnaire, and a fall history, which adds time and burden for patients and clinicians.

Because the conditions share motor impairments, 1 motor assessment could screen for all 3, and sensor-based technologies can measure motor function precisely and at low cost. Depth cameras, force plates, and wearable sensors can quantify gait, balance, sit-to-stand performance, and reaction time, including under dual-task conditions in which a motor task is performed together with a cognitive task [14,15]; dual-task gait in particular has been linked to cognitive impairment and progression to dementia [16,17]. Machine learning (ML) models built on such measurements have shown promise for identifying MCI [14], frailty [18,19], and fall risk [15,20–23]. However, each of these models predicts 1 condition, so it cannot use the information the conditions share, and a separate model must be built and validated for each outcome. A unified model removes this limitation. A classifier chain predicts several outcomes with a unified model by using the prediction for each outcome as an input for the next, so that the relationships between outcomes can be used [24].

However, 3 gaps remain before such a model can support screening. First, to our knowledge, no study has developed a unified model for MCI, frailty, and fall risk and compared it directly with separate models under the same validation settings. The few unified models in health care predicted other outcomes from other data, such as diabetic complications from electronic health records [25], and frailty-related outcomes from administrative data [26]. In both, the unified model outperformed separate models, but neither used sensor-based motor-function data or predicted cognitive and physical decline together. Second, sensor-based studies usually include fewer than 100 participants and evaluate their models on a single small held-out set. Such estimates depend on which participants are tested, can overstate performance on new individuals, and are rarely reported with a measure of uncertainty [27,28]. Third, Shapley additive explanations (SHAP) are widely used to explain model predictions [29], but these explanations are rarely tested on unseen participants or compared with actual differences between people with and without the condition. Without this evidence, it is difficult to judge whether a model can be trusted in clinical practice [30].

To address these gaps, this study aimed to develop an explainable unified ML model that predicts MCI, frailty, and fall risk together from the multimodal motor-function, demographic, and family-history data of community-dwelling older adults. To obtain consistent estimates in a small sample, we evaluated the model with repeated nested cross-validation and reported 95% CIs. The principal contributions of this study are as follows:

1. We propose an explainable unified ML model that predicts MCI, frailty, and fall risk together from a single multimodal motor-function assessment, and we compare it directly with single-label models developed under the same settings.
2. We provide SHAP explanations for each outcome that separate shared from condition-specific features. We test whether these explanations are consistent between training and unseen participants in the outer test folds, and link the top features to differences between participants with and without each condition.
3. We test whether the classifier chain uses the dependencies between conditions by shuffling the predicted conditions used as inputs in the chain. We also test its robustness to the chain order and to the removal of each assessment domain.

## Methods

### Study Overview

We used cross-sectional data from community-dwelling older adults who completed a multimodal motor-function assessment to test whether a unified model can predict MCI, frailty, and fall risk from 1 assessment. We developed a unified multilabel model that predicts the 3 conditions together and compared it with single-label models that predict each condition separately. Both approaches used the same participants, features, algorithms, cross-validation splits, and evaluation methods, so they differed only in whether the 3 outcomes were predicted jointly or separately (Figure 1).

**Figure 1.**
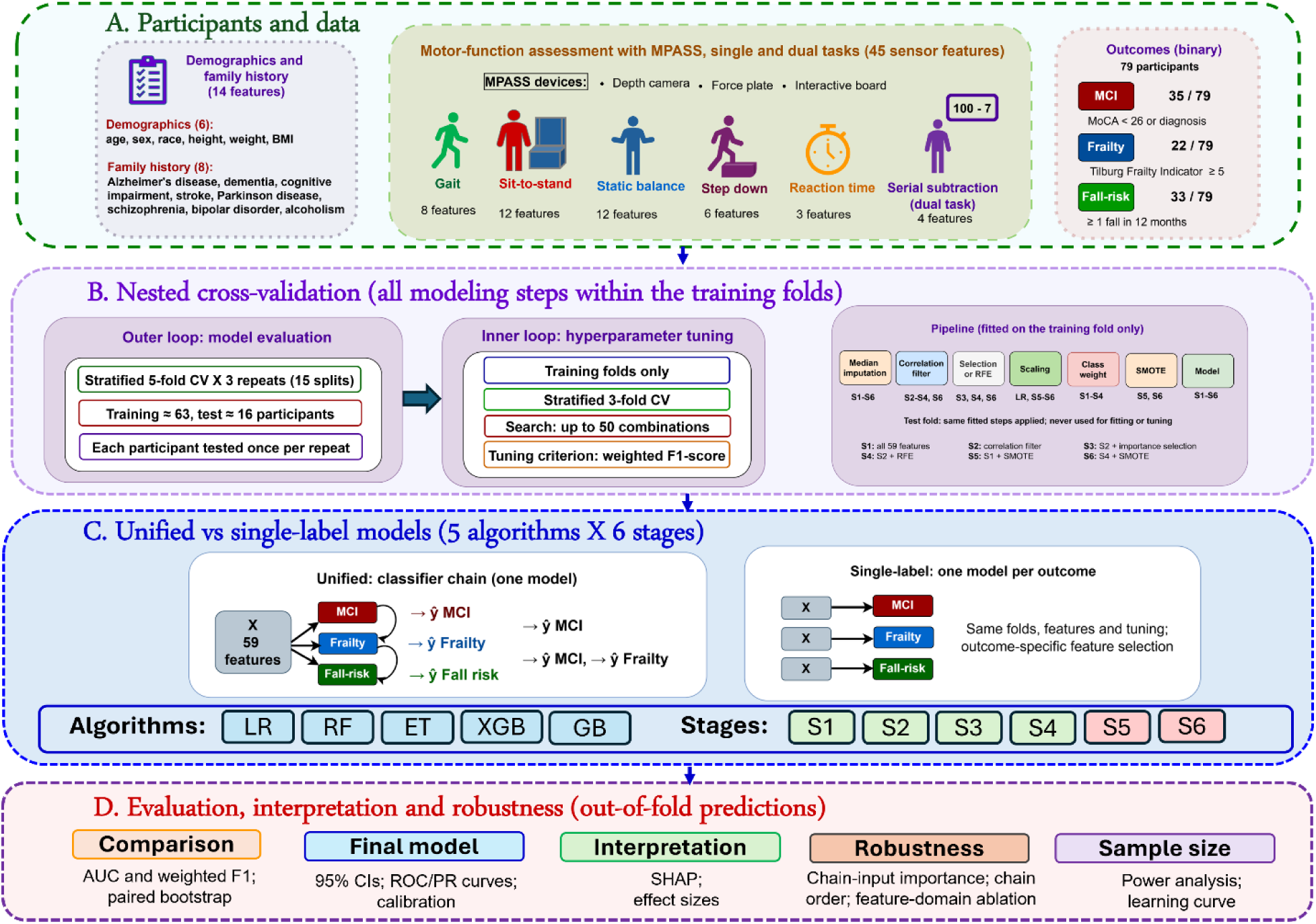
Analytical workflow. (A) Participants, features, and outcomes. (B) Nested cross-validation, with all data-dependent steps fitted on the training folds only. (C) Unified and single-label models compared across 5 algorithms and 6 modeling stages. (D) Evaluation, interpretation, and robustness analyses based on out-of-fold predictions.

### Participants and Data

Adults aged 60 years and older were recruited from the University of Missouri Health Psychology Clinic and the surrounding mid-Missouri communities. Eligible participants could walk independently without assistive devices and had no orthopedic or neurological condition that could affect movement. Of the 83 enrolled participants, 4 had missing outcome data and were excluded, leaving 79 for analysis.

In a single visit, each participant completed a demographic and health-history questionnaire, the Montreal Cognitive Assessment (MoCA), and 5 motor-function tasks recorded with the Mizzou Point-of-Care Assessment System (MPASS), a portable platform developed at the University of Missouri that combines a depth camera, a custom-built force plate, and an interactive interface board [14,31,32]. Participants walked a 6-m path 3 times (gait), rose from a 44-cm bench 5 times with arms crossed (sit-to-stand [STS]), stood quietly on the force plate (static balance), stepped down from a 21-cm platform 3 times with each leg (step-down), and responded to randomly presented red and green light cues (reaction time). The gait, STS, and static-balance tasks were repeated under a dual-task condition, in which participants performed serial subtraction (counting backward by 7s). Separate models for each condition were previously developed with this cohort [31]; this study tests whether MCI, frailty, and fall risk can be predicted together with a unified model.

### Ethical Considerations

The study was approved by the University of Missouri Institutional Review Board (IRB No. 2102007). All participants provided written informed consent before data collection and received a gift card as compensation. Data were de-identified before analysis.

### Outcome Definitions

MCI was defined as a MoCA score below 26 or a clinical diagnosis by a board-certified neuropsychologist [33], frailty as a Tilburg Frailty Indicator score of 5 or higher [34], and fall risk as 1 or more self-reported falls in the previous 12 months. Fall history was used as an indicator of fall risk, because a previous fall is one of the strongest predictors of future falls [6]. Participants who met a criterion were coded as cases (1) and all others as controls (0). Because the outcomes are not mutually exclusive, a participant could be a case for more than one outcome.

### Features and Data Preprocessing

The depth camera (30 Hz) captured the center of mass and foot placement during gait, knee movement during STS, and the center of mass during step-down; its gait and STS measurements have been validated [35,36]. The force plate recorded center-of-pressure (CoP) movement during static balance, and the interface board recorded button responses in the reaction-time task. These recordings were processed in MATLAB (MathWorks) to extract 45 sensor features, which were averaged over repeated trials and combined with 6 demographic and 8 self-reported family-history variables, giving 59 features in 8 domains (Table 1; Multimedia Appendix 1, Table S1). Sex (0=male, 1=female), race (0=White, 1=non-White), and each family history (0=no, 1=yes) were coded as binary variables.

**Table 1.** Features used in the models, by domain.

| **Domain** | **Source** | **Features, n** | **Measures** |
|---|---|---|---|
| Gait | Depth camera | 8 | Stride length and stride time, left and right; ST and DT |
| Sit-to-stand | Depth camera | 12 | Completion time and knee angular velocity while rising and sitting, over 3 and 5 repetitions; ST and DT |
| Static balance | Force plate | 12 | CoP 95% sway area; mean CoP velocity (total, ML, and AP); AUC (ML and AP); ST and DT |
| Step-down | Depth camera | 6 | Leading- and trailing-limb time and ML center-of-mass displacement, left and right |
| Reaction time | Interface board | 3 | Reaction time, incorrect presses, and buttons pressed |
| Serial subtraction | Static-balance DT trials | 4 | Total and incorrect responses in 15 s and dual-task interference (total and correct); DT |
| Demographics | Questionnaire | 6 | Age, sex, height, weight, BMI, and race |
| Family history | Questionnaire | 8 | Alzheimer disease, dementia, Parkinson disease, stroke, cognitive impairment, schizophrenia, bipolar disorder, and alcoholism |
| **Total** |  | **59** |  |

Sensor features were averaged over repeated trials. Full feature names and descriptions are given in Multimedia Appendix 1 (Table S1). AP: anteroposterior; CoP: center of pressure; DT: dual task; ML: mediolateral; ST: single task.

Of all feature values, 149 (3.2%) were missing. Missing values were replaced with the median of the training participants in each cross-validation fold, because sensor measures are often skewed. Features were standardized with the mean and SD of the training participants for logistic regression, whose regularization depends on feature scale, and before oversampling, which uses distances between participants. The tree-based algorithms used unscaled features, because their splits do not depend on feature scale.

### Nested Cross-Validation and Hyperparameter Tuning

All models were developed and evaluated with repeated nested cross-validation, which tests every participant and keeps hyperparameter tuning separate from performance evaluation [27]. With 79 participants, one held-out test set would have been too small for reliable estimates: an 80/20 split would have tested only 16 participants, including 4 or 5 with frailty, and the results would have depended heavily on which participants were tested [28].

The outer loop estimated performance. The 79 participants were divided into 5 folds (4 of 16 participants and 1 of 15), stratified by outcome combination, with combinations seen in fewer than 5 participants grouped together. Each fold served once as the test fold, and the other 63 or 64 participants formed the training set. Repeating this 3 times with new random folds gave 15 train-test splits, in which each participant was tested 3 times, always by a model not trained on them. The same 15 splits were used for all models and analyses, so the unified and single-label models could be compared on the same participants.

The inner loop tuned the hyperparameters within each outer training set, which was divided into 3 stratified folds; each hyperparameter combination was trained on 2 folds and evaluated on the third, in turn. Every algorithm had the same tuning budget: all combinations when the search space contained 50 or fewer, and 50 randomly sampled combinations otherwise [37] (Multimedia Appendix 2, Table S2). In S3, S4, and S6, the feature-selection settings were tuned together with the hyperparameters. The combination with the highest mean weighted F1-score was refitted on the full outer training set and used to predict the outer test fold. The weighted F1-score weighted cases and controls by their numbers for the single-label models, and the 3 outcomes by their numbers of cases for the unified model.

To prevent information from the test participants from entering model development, all data-dependent steps (median imputation, correlation filtering, feature selection, standardization, oversampling, and model fitting) were fitted on the training participants only and applied unchanged to the test participants, in both loops. A fixed random seed (42) was used for all splits, searches, and models.

### Unified and Single-Label Models

We compared 2 approaches: a unified multilabel model that predicts MCI, frailty, and fall risk jointly, and single-label models that predict each outcome separately.

The unified model used a classifier chain, in which each outcome is used as an extra input to the classifiers of the following outcomes [24]. With the outcomes ordered as MCI, frailty, and fall risk, the joint probability of the 3 outcomes is:

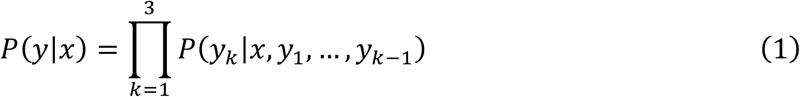

where x denotes the feature vector, yₖ denotes outcome k in chain order (k=1, 2, 3 for MCI, frailty, and fall risk, respectively), and y₁,…,yₖ₋₁ denote the preceding outcomes. All 3 classifiers used the 59 features. The frailty classifier also received the MCI label, and the fall-risk classifier received both the MCI and frailty labels. During training, these extra inputs were the observed labels (y₁,…,yₖ₋₁); for test participants, they were the labels predicted in the earlier steps of the chain (ŷ₁,…,ŷₖ₋₁). The chain order was chosen because cognitive impairment and frailty together increase the risk of falls [8]. Because the 3 conditions were measured at the same visit, this order is a modeling choice rather than a description of disease progression, and its effect was examined in a sensitivity analysis of all 6 possible orders. In the unified model, the 3 outcomes shared a single set of features and hyperparameters.

The single-label approach fitted a separate classifier for each outcome:

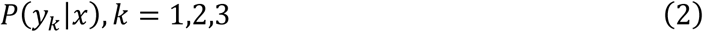

Each single-label classifier was tuned separately, selected its own features when feature selection was applied, and shared no information with the other outcomes.

Both approaches were implemented with 5 algorithms covering linear, tree-ensemble, and boosting methods. Logistic regression (LR) models the log-odds of each outcome as a linear combination of the features, with L1 or L2 regularization [38]. Random forest (RF) [39] and extra trees (ET) [40] are ensembles of decision trees that consider a random subset of features at each split. RF trains each tree on a bootstrap sample of participants, whereas ET chooses split thresholds at random, which further reduces variance; whether ET used bootstrap samples was tuned as a hyperparameter. Gradient boosting (GB) [41] and extreme gradient boosting (XGBoost) [42] build trees sequentially, with each tree correcting the errors of the previous ones.

### Feature-Selection and Class-Imbalance Strategies

To test whether feature selection or synthetic oversampling improved on using all features with class weighting, we evaluated 6 modeling stages (Table 2). In S3 and S4, the unified model trained the selection model on all 3 outcomes together and selected 1 shared feature set, whereas each single-label model selected its own features for its outcome. All steps were fitted on the training data only.

- **All features (S1):** All 59 features were used with class weights, and S1 served as the reference.
- **S2 (correlation filter):** Highly correlated features were removed. Pearson correlations between all feature pairs were calculated, and pairs with an absolute correlation above 0.90 were processed from the highest correlation downward. In each pair, the feature with the higher mean absolute correlation with all other features was removed, and pairs with an already removed feature were skipped. Applied to all 79 participants, the filter removed 14 features, leaving 45; all highly correlated pairs (|r|=0.91-0.99) were within the same domain, such as left and right stride length or the 3- and 5-repetition STS measures (Multimedia Appendix 3, Figure S1).
- **S3 (correlation filter + importance selection):** After the correlation filter, an ET model (300 trees, balanced class weights) ranked the remaining features by mean decrease in Gini impurity, and features at or above the median or the mean importance were kept, with the threshold tuned during cross-validation.
- **S4 (correlation filter + recursive feature elimination [RFE]):** After the correlation filter, an ET model (100 trees, balanced class weights) was fitted repeatedly, each time removing the 20% of remaining features with the lowest importance, until the target number of features (5 to 45, in steps of 5, tuned during cross-validation) remained.
- **S5 (all features + SMOTE).** Same features as S1, but class weights were replaced with the synthetic minority oversampling technique (SMOTE) [43].
- **S6 (correlation filter + RFE + SMOTE):** Same feature selection as S4, but class weights were replaced with SMOTE, applied after feature selection so that features were selected using real participants only.

**Table 2.** Modeling stages evaluated for the unified and single-label models.

| **Stage** | **Feature selection** | **Class imbalance** | **Number of features** |
|---|---|---|---|
| S1 | None | Class weights | 59 |
| S2 | Correlation filter (\|r\| > 0.90) | Class weights | 45<sup>a</sup> |
| S3 | Correlation filter + importance selection | Class weights | Tuned (median or mean threshold) |
| S4 | Correlation filter + RFE | Class weights | Tuned (5–45) |
| S5 | None | SMOTE | 59 |
| S6 | Correlation filter + RFE | SMOTE | Tuned (5–45) |

<sup>a</sup>Number of features retained when the correlation filter was applied to all 79 participants; because the filter was refitted within each training set, the number could differ slightly between splits.

### Performance Evaluation and Statistical Comparison

Because each outer test fold contained only 15 or 16 participants, predictions were pooled: in each repeat, the 5 test folds together gave a single out-of-fold prediction for each of the 79 participants. Metrics were calculated from these 79 predictions and averaged over the 3 repeats.

The primary metric was the area under the receiver operating characteristic (ROC) curve (AUC), because it does not depend on a classification threshold or on the prevalence of the outcome. The secondary metric was the weighted F1-score, which was also used for tuning and reflects classification at a threshold of 0.5. We also calculated the area under the precision-recall curve (PR-AUC) [44], whose chance level equals the prevalence, and, at the same threshold, sensitivity, specificity, positive predictive value (PPV), negative predictive value (NPV), and accuracy. All metrics were calculated for each outcome and averaged across the 3 outcomes.

The unified and single-label models were compared with a paired bootstrap [45]. In each of 1000 bootstrap samples, 79 participants were drawn with replacement, both approaches were evaluated on the same participants, and the difference (unified − single-label) was averaged over the 3 repeats; samples in which an outcome had only a single class were excluded. The 95% CI was taken from the 2.5th and 97.5th percentiles of the differences, the 2-sided P value as twice the proportion of differences on the less frequent side of 0, and a difference was considered significant when the 95% CI excluded 0. The same bootstrap was used for all other CIs and comparisons in the study.

### Final Model Selection

We tested 30 unified models (5 algorithms × 6 stages) and chose the one with the highest mean AUC as the final model, using the weighted F1-score, accuracy, and PR-AUC to decide between near ties. Because choosing the best of many models on the same test predictions can overstate performance, we also made the selection inside the cross-validation (nested model selection): in each outer training set, all 20 class-weighted models (S1-S4) were tuned, and only the one with the highest inner weighted F1-score predicted the outer test fold. The unified approach chose a single algorithm and stage for all 3 outcomes, whereas the single-label approach chose one for each outcome, so this test performance includes the effect of model selection.

For the final model, ROC and precision-recall curves and confusion matrices were built from each participant’s predicted probabilities averaged over the 3 repeats, at a threshold of 0.5. Performance across the 3 outcomes together was summarized with subset accuracy, the proportion of participants with all 3 outcomes predicted correctly.

Calibration, the agreement between predicted and observed risk, was assessed with the Brier score [46], compared with the score obtained by predicting the prevalence for every participant (prevalence × [1 − prevalence]); with the calibration intercept and slope (ideal values 0 and 1), estimated by logistic regression of the observed outcome on the logit of the predicted probability; and by comparing observed and predicted risks within quintiles of predicted risk.

Four sensitivity analyses tested whether these results depended on choices in the main analysis:

- Extended LR grid: LR with 59 features and about 63 training participants may need stronger regularization, so it was repeated with C=0.001, 0.003, 0.01, 0.03, 0.1, 1, 10, and 100.
- AUC tuning: because the weighted F1-score depends on the threshold, S1 was repeated with hyperparameters tuned for AUC.
- No class weights: class weighting can improve detection of the smaller group but reduce calibration [47], so S1 was repeated without class weights.
- Probabilities in the chain: because predicted labels (0 or 1) discard the uncertainty of each prediction, the unified S1 models were repeated with predicted probabilities as chain inputs.

### Model Interpretation

The predictions of the final model were explained with Shapley additive explanations (SHAP), which assign each feature a contribution to each prediction that moves the predicted probability toward or away from the condition [29]. SHAP values were calculated on the probability scale with TreeExplainer [48] for each step of the chain in each of the 15 outer splits, for both the training and the test participants, and averaged per participant across splits; the test values show whether the explanations hold for unseen participants.

For each feature, we calculated the mean absolute SHAP value for each outcome in the training set. A feature’s importance was the sum of these values over the 3 outcomes, and each outcome’s share of it showed which outcomes the feature served. A feature was considered shared unless a single outcome accounted for 60% or more of its importance. A feature was considered shared unless a single outcome accounted for 60% or more of its importance. The predicted outcomes used as chain inputs were excluded because they are not inputs of every step. The Spearman rank correlation measured the similarity of the training and test rankings. The beeswarm plots average each participant’s SHAP values across the 3 outcomes; for demographic and family-history features, the direction was also calculated for each outcome separately. SHAP results were reported for the test participants, with features ordered by training-set importance, and separately for the training participants.

### Effect-Size Analysis

To test whether the most important features differed between participants with and without each condition, we calculated standardized mean differences (SMDs) for all 59 features in all 79 participants, using median-imputed values: Hedges g for continuous features [49] and the SMD for proportions for binary features [50]:

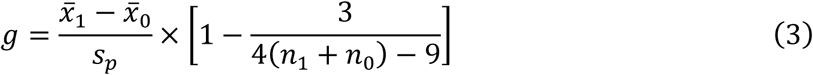

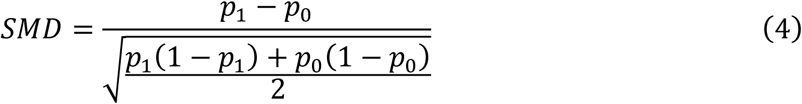

where x̄₁ and x̄₀ are the group means, sₚ the pooled SD, n₁ and n₀ the group sizes, and p₁ and p₀ the proportions with the feature in participants with and without the condition; the bracket corrects g for small samples. Absolute SMDs of 0.2, 0.5, and 0.8 indicated small, medium, and large effects [51]. For the 10 most important features, group differences were tested with the Mann-Whitney U test (continuous features) or the Fisher exact test (binary features), with Benjamini-Hochberg correction [52]. Because these analyses used all participants, they were descriptive only and were not used for model development.

### Robustness Analyses

Three analyses examined the robustness of the final model. All used the same 15 splits, tuning procedure, and test participants as the main analysis, so any change in performance could be attributed to the factor tested, and changes in AUC were compared with the paired bootstrap.

#### Chain-Input Importance

To test whether the model used the predicted outcomes passed along the chain, each input of the frailty and fall-risk steps (the 59 features and the chain inputs) was shuffled 20 times among the test participants in each split. The decrease in test AUC was computed from the pooled test predictions of each repeat and averaged over the 3 repeats. Each chain input was ranked among all inputs of its step (60 in the frailty step and 61 in the fall-risk step) and compared with the most important original feature. We used permutation importance on the outer test folds instead of Gini importance, because Gini importance is calculated on the training data and underestimates binary inputs such as the chain inputs [53].

#### Feature-Domain Ablation

To test whether the model depended on any single type of assessment, each of the 8 domains was removed in turn, and the model was retrained. A ninth model used only the 45 sensor features, without demographics and family history, to evaluate the motor-function assessment alone.

#### Chain-Order Sensitivity

Because the order determines which predictions are available at each step, the model was retrained with all 6 possible orders of the outcomes, and the AUC of each outcome and the mean AUC were compared with the main order.

### Sample-Size Analyses

#### Learning Curve

To test whether performance had reached a plateau or would improve with more participants [54], the model was retrained on 40%, 55%, 70%, 85%, and 100% of each outer training set (on average 25, 34, 44, 53, and 63 participants). Each reduced set was sampled at random and stratified by outcome combination, with at least 3 cases and 3 controls for each outcome so that inner 3-fold tuning remained possible. Each model was tuned as in the main analysis and tested on the same test participants. Test AUC at each size was compared with the full size, and training AUC was recorded to assess overfitting.

#### Sensitivity Power Analysis

To estimate the smallest group difference the sample could detect, we used the observed group sizes for each outcome (35 vs 44 for MCI, 22 vs 57 for frailty, and 33 vs 46 for fall risk) and a 2-sided significance level of.05. We calculated the smallest SMD detectable with 80% power, the power to detect small, medium, and large effects, and the sample size needed to detect each with 80% power. Power based on the observed effect sizes was not calculated, because it adds no information beyond the P value [55].

### Software

Sensor data were processed in MATLAB, and all other analyses were conducted in Python 3.9.13 with scikit-learn 1.6.1, XGBoost 2.1.4, SHAP 0.47.2, SciPy 1.11.4, NumPy 1.26.4, pandas 2.0.3, and Matplotlib 3.9.4. SMOTE was implemented directly so that it could oversample the outcome combinations of the unified model and be applied only to the training data.

### Results

We first compared the unified model with separate single-label models, then evaluated the final model, and finally examined how it made its predictions and whether it used the links between the conditions.

### Participant Characteristics

The 79 participants had a mean age of 70.86 (SD 4.68) years; 48 (60.8%) were female, and 77 (97.5%) were White (Table 3). The 3 conditions were common in this community sample: MCI affected 35 (44.3%) participants, fall risk 33 (41.8%), and frailty 22 (27.8%). The most frequent family histories were stroke (35.4%), dementia (34.2%), and alcoholism (29.1%). Participants with MCI had a higher weight (84.40 vs 75.57 kg; P=.02) and BMI (29.81 vs 26.53 kg/m²; P=.006), whereas no characteristic differed significantly by frailty or fall risk (Multimedia Appendix 4, Table S3).

**Table 3.**
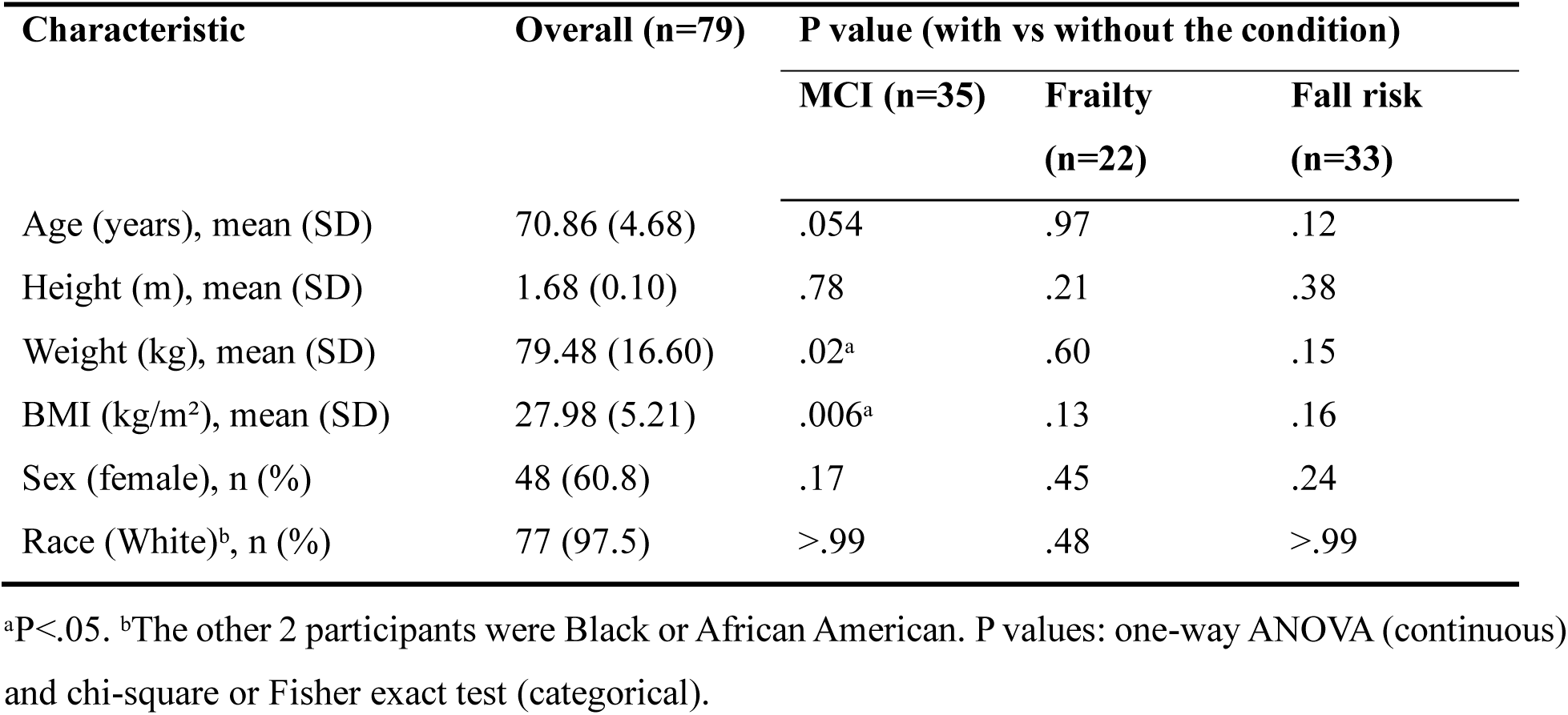
Characteristics of the participants (N=79) and comparison between participants with and without each condition.

### Unified and Single-Label Approaches

With class weighting, the unified model achieved a higher mean AUC than the single-label models in 17 of 20 configurations and was never significantly worse (Table 4). Both significant differences showed a higher AUC for the unified model: XGBoost in S3 (+0.031, 95% CI 0.001 to 0.061; P=.048) and ET in S4 (+0.050, 95% CI 0.013 to 0.089; P=.01). The unified model was also more robust to feature selection: from S1 to S4, its best mean AUC decreased only from 0.645 to 0.633, whereas that of the single-label models decreased from 0.648 to 0.607 (Figure 2; Multimedia Appendix 4, Table S4). With all features (S1), the best unified models (ET and XGBoost) reached a mean AUC of 0.645 and the best single-label model (RF) 0.648, and both approaches predicted MCI best (unified AUC 0.733-0.781; single-label 0.722-0.789), followed by frailty (0.560-0.630 vs 0.480-0.622) and fall risk (0.485-0.572 vs 0.465-0.580; Multimedia Appendix 4, Table S5).

**Figure 2.**
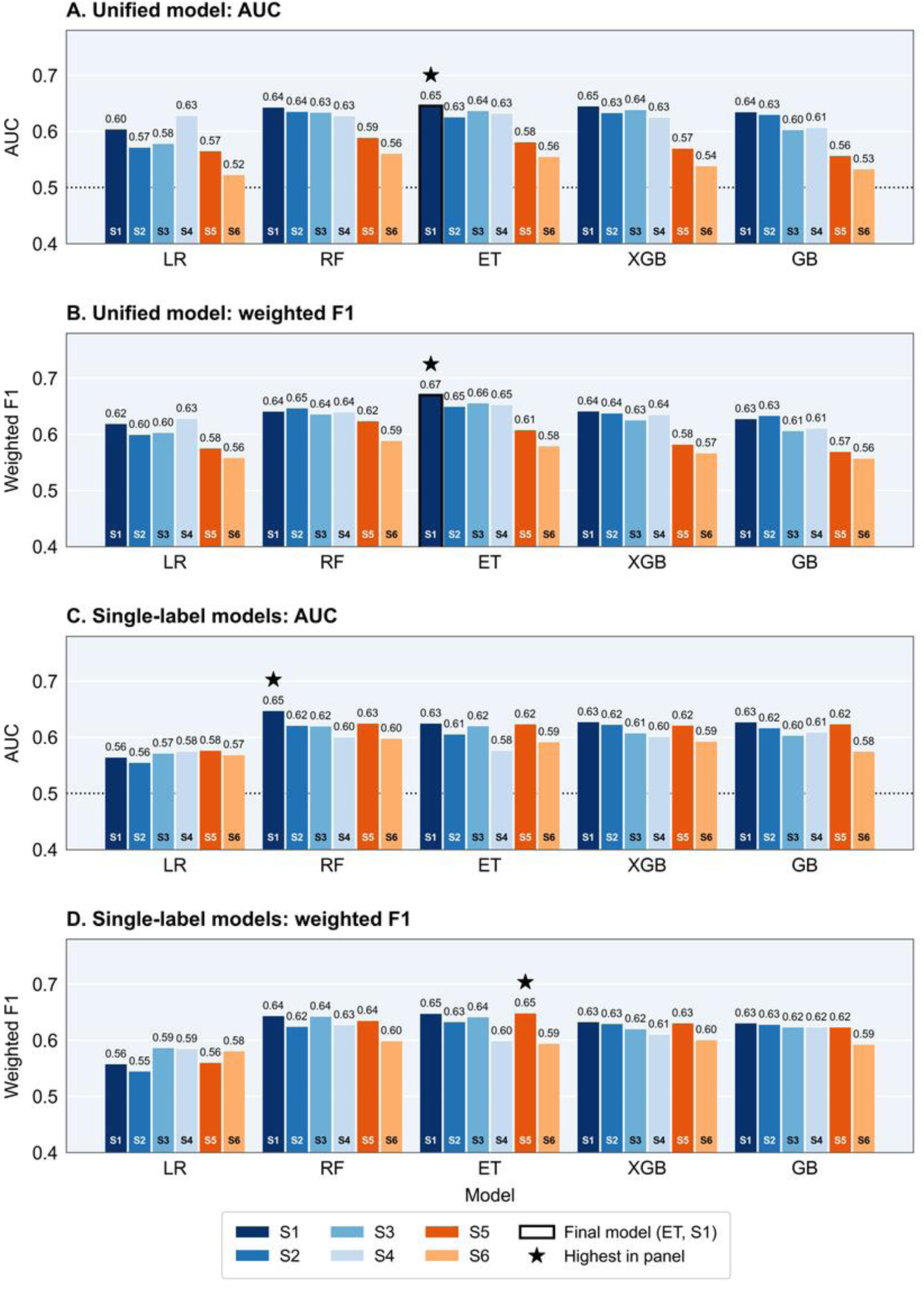
Mean AUC and weighted F1-score of the unified and single-label models by algorithm and modeling stage. (A, B) Unified and (C, D) single-label models. Values are means over the 3 outcomes. Blue bars: class weights (S1-S4); orange bars: SMOTE (S5-S6). Outlined bar: final model (ET, S1); star: highest value in each panel; dotted line (A, C): chance (AUC 0.5).

**Table 4.** Comparison of the unified and single-label models by modeling stage.

| **Stage** | **Best unified mean AUC (algorithm)** | **Best single-label mean AUC (algorithm)** | **Algorithms with higher unified AUC** | **Significant differences in mean AUC, unified − single-label (95% CI)** |
|---|---|---|---|---|
| S1 | 0.645 (ET) | 0.648 (RF) | 4/5 | — |
| S2 | 0.636 (RF) | 0.623 (XGBoost) | 5/5 | — |
| S3 | 0.639 (XGBoost) | 0.620 (RF, ET) | 4/5 | XGBoost: +0.031 (0.001 to 0.061); *P*=.048 |
| S4 | 0.633 (ET) | 0.607 (XGBoost) | 4/5 | ET: +0.050 (0.013 to 0.089); *P*=.01 |
| S5 | 0.589 (RF) | 0.625 (RF) | 0/5 | GB: −0.067 (−0.122 to −0.015); *P*=.01 |
|  |  |  |  | XGBoost: −0.051 (−0.091 to −0.016); *P*=.008 |
|  |  |  |  | ET: −0.042 (−0.086 to −0.005); *P*=.03 |
| S6 | 0.559 (RF) | 0.605 (RF) | 0/5 | LR: −0.051 (−0.096 to −0.003); *P*=.03 |

Mean AUC across the 3 outcomes from pooled out-of-fold predictions. Differences, 95% CIs, and *P* values from the paired bootstrap; positive values indicate a higher AUC for the unified model; −, no significant difference. S1-S4 used class weights, and S5-S6 used SMOTE (line). Full results in Multimedia Appendix 4 (Table S4).

The unified model gained most where single-label models struggled: frailty and fall risk, the 2 harder-to-predict outcomes (Figure 3). In S1, it had a significantly higher frailty AUC with LR (+0.081; P=.01) and GB (+0.061; P=.048), and in S4 its largest gains were again for frailty and fall risk (ET: +0.061 and +0.070; P=.06 and P=.10). For MCI, the 2 approaches performed similarly in every stage.

**Figure 3.**
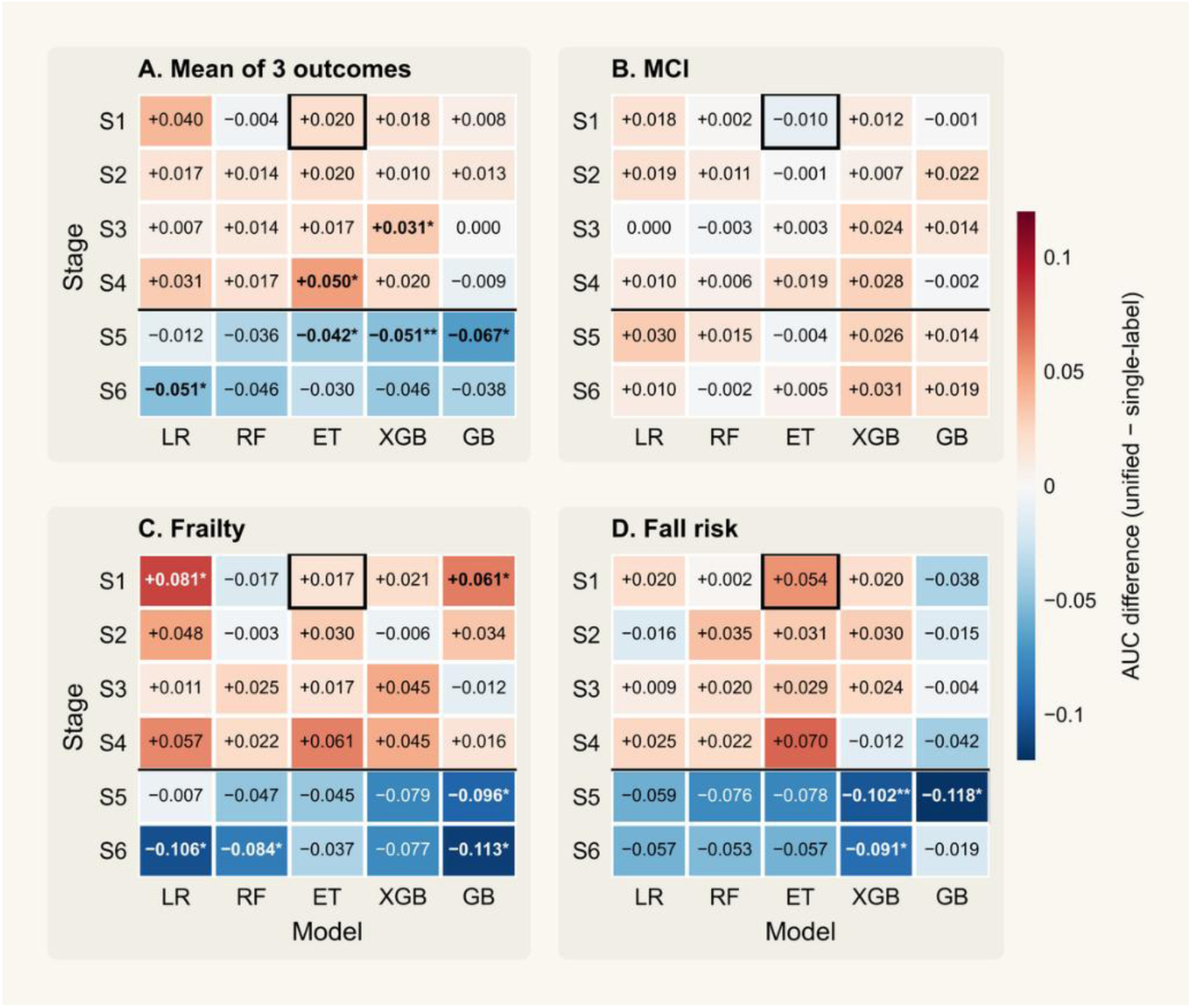
Difference in test AUC between the unified and single-label models by algorithm and modeling stage. (A) Mean, (B) MCI, (C) frailty, and (D) fall risk. Positive values favor the unified model; the line separates class weighting (S1-S4) from SMOTE (S5-S6); outlined cell: final model. *P<.05, **P<.01 (paired bootstrap).

This advantage depended on how class imbalance was handled. With SMOTE (S5-S6), the unified model had a lower mean AUC in all 10 combinations, and all 4 significant differences showed a higher AUC for the single-label models (Table 4); the best unified mean AUC decreased to 0.589 in S5 and 0.559 in S6, compared with 0.625 and 0.605 for the single-label models. All later analyses therefore used class weighting.

The comparison held across tuning and selection choices (Multimedia Appendix 4, Tables S6 and S7). A wider LR regularization range changed its mean AUC by ≤0.016. Tuning for AUC instead of weighted F1-score changed each model’s mean AUC by ≤0.011, except single-label LR (0.565 to 0.598), and unified ET remained the best unified model (0.653). Removing class weights (differences −0.011 to +0.013; all P≥.34) or passing predicted probabilities along the chain (−0.027 to 0.000; all P≥.052) did not change the comparison. When the algorithm and stage were also chosen inside the cross-validation (nested model selection), the unified model again performed at least as well as the single-label models (mean AUC 0.615 vs 0.605; difference +0.011, 95% CI−0.034 to 0.051; P=.58; weighted F1-score 0.622 for both).

### Performance of the Final Model

The final unified model (ET, all features, S1) achieved a mean AUC of 0.645 (95% CI 0.569-0.722) (Table 5; Figure 4A). It tied with XGBoost on mean AUC and had the higher weighted F1-score (Figure 2B) and PR-AUC (Multimedia Appendix 4, Table S5). Discrimination was best for MCI (AUC 0.779, 95% CI 0.680-0.872) and lower for frailty (0.606) and fall risk (0.551); only for MCI did the 95% CIs of AUC and PR-AUC lie entirely above chance (Figure 4B).

**Figure 4.**
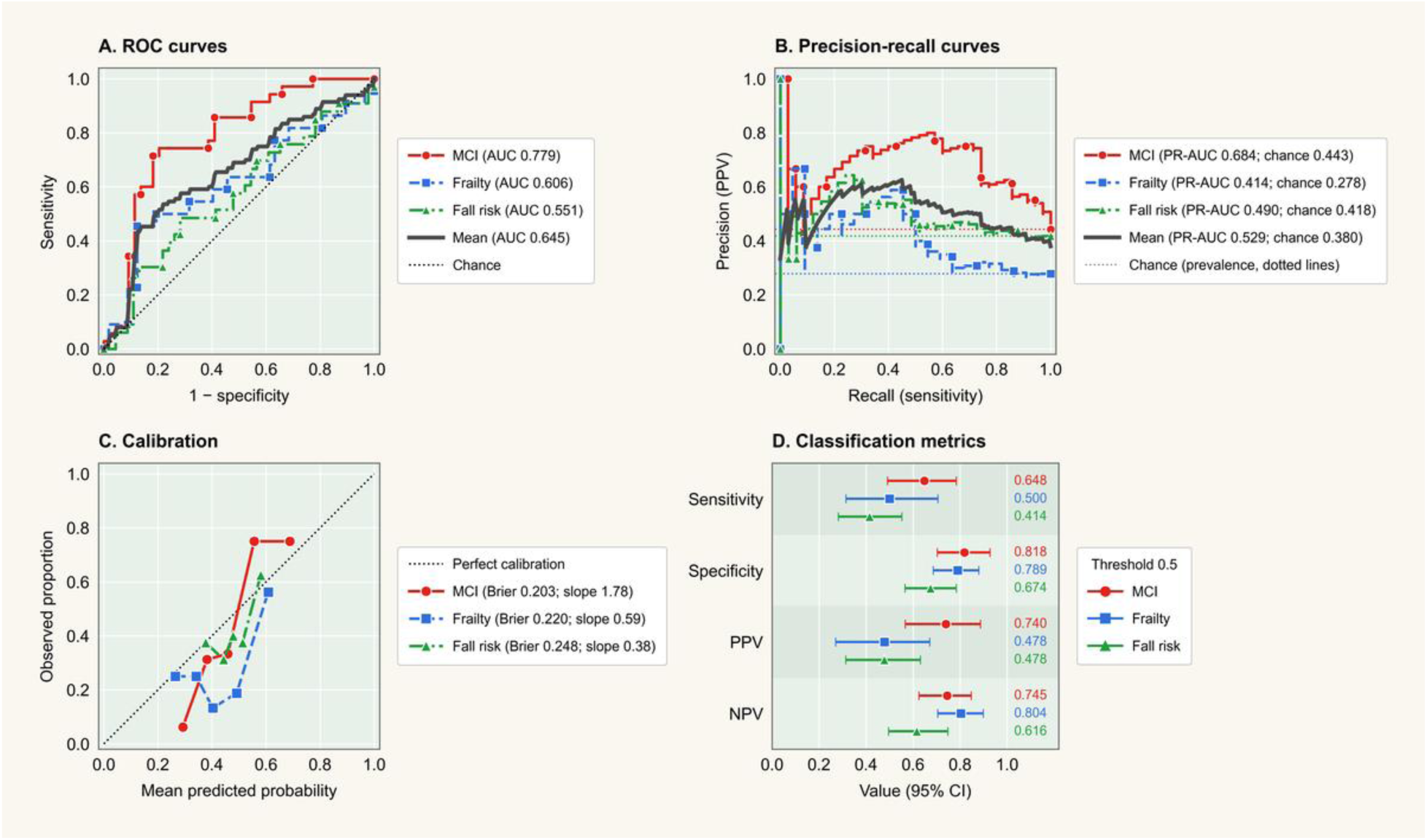
Discrimination, calibration, and classification performance of the final unified model for each outcome. (A) ROC and (B) precision-recall curves; dotted lines mark chance (in B, the prevalence). (C) Calibration within quintiles of predicted risk; dotted line: perfect calibration. (D) Sensitivity, specificity, PPV, and NPV (95% CI) at a threshold of 0.5.

**Table 5.**
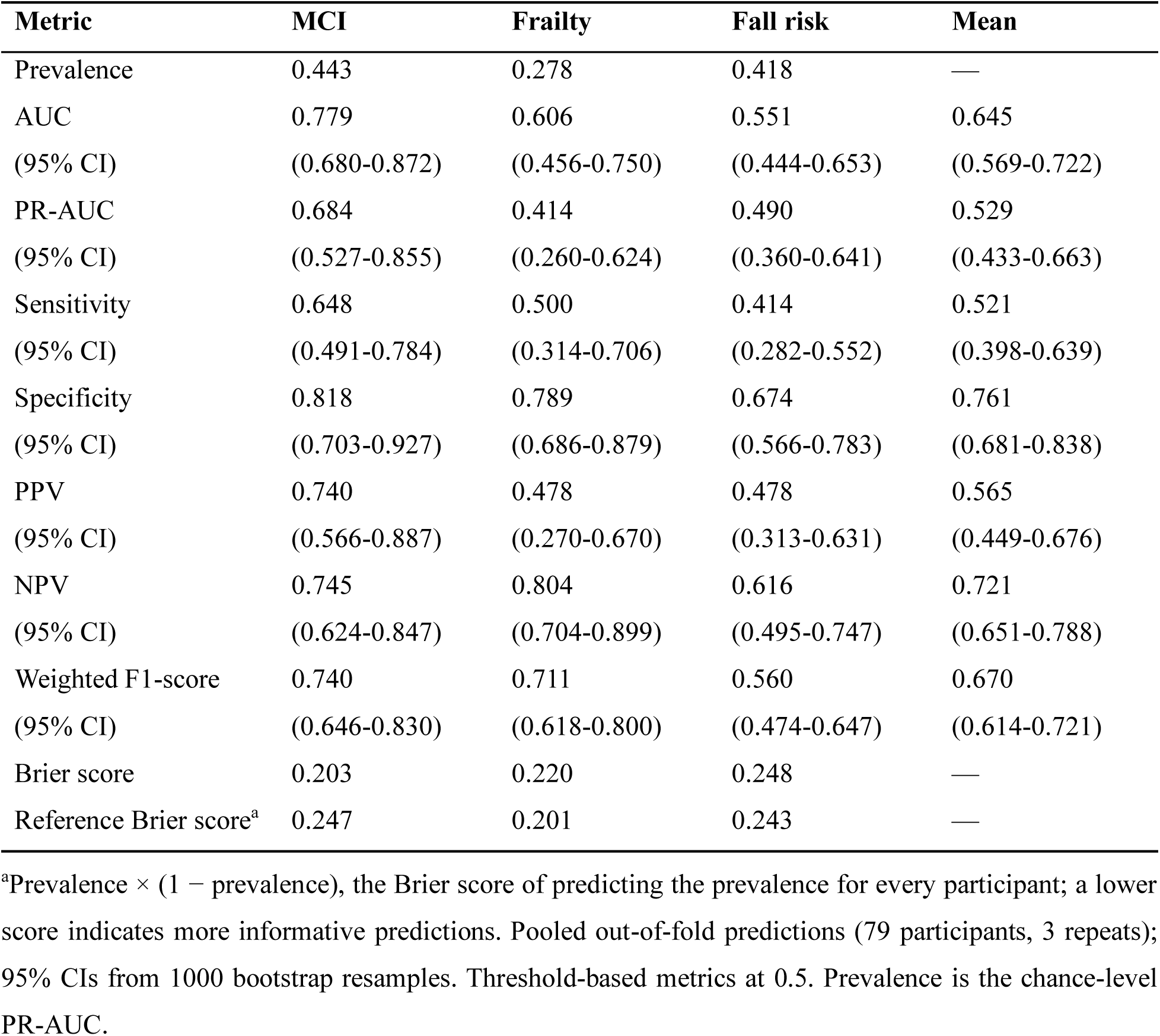
Performance and calibration of the final unified model (extra trees, all features) for each outcome.

| **Metric** | **MCI** | **Frailty** | **Fall risk** | **Mean** |
|---|---|---|---|---|
| Prevalence | 0.443 | 0.278 | 0.418 | — |
| AUC | 0.779 | 0.606 | 0.551 | 0.645 |
| (95% CI) | (0.680-0.872) | (0.456-0.750) | (0.444-0.653) | (0.569-0.722) |
| PR-AUC | 0.684 | 0.414 | 0.490 | 0.529 |
| (95% CI) | (0.527-0.855) | (0.260-0.624) | (0.360-0.641) | (0.433-0.663) |
| Sensitivity | 0.648 | 0.500 | 0.414 | 0.521 |
| (95% CI) | (0.491-0.784) | (0.314-0.706) | (0.282-0.552) | (0.398-0.639) |
| Specificity | 0.818 | 0.789 | 0.674 | 0.761 |
| (95% CI) | (0.703-0.927) | (0.686-0.879) | (0.566-0.783) | (0.681-0.838) |
| PPV | 0.740 | 0.478 | 0.478 | 0.565 |
| (95% CI) | (0.566-0.887) | (0.270-0.670) | (0.313-0.631) | (0.449-0.676) |
| NPV | 0.745 | 0.804 | 0.616 | 0.721 |
| (95% CI) | (0.624-0.847) | (0.704-0.899) | (0.495-0.747) | (0.651-0.788) |
| Weighted F1-score | 0.740 | 0.711 | 0.560 | 0.670 |
| (95% CI) | (0.646-0.830) | (0.618-0.800) | (0.474-0.647) | (0.614-0.721) |
| Brier score | 0.203 | 0.220 | 0.248 | — |
| Reference Brier score<sup>a</sup> | 0.247 | 0.201 | 0.243 | — |

<sup>a</sup>Prevalence × (1 − prevalence), the Brier score of predicting the prevalence for every participant; a lower score indicates more informative predictions. Pooled out-of-fold predictions (79 participants, 3 repeats); 95% CIs from 1000 bootstrap resamples. Threshold-based metrics at 0.5. Prevalence is the chance-level PR-AUC.

At a threshold of 0.5, specificity was higher than sensitivity for all 3 outcomes, so the model identified participants without a condition more reliably than those with it (Figure 4D). For MCI, it identified 64.8% of participants with MCI and 81.8% of those without, and 74.0% of its positive predictions were correct. For frailty, negative predictions were usually correct (NPV 0.804), but fewer than half of positive predictions were (PPV 0.478). Fall risk had the lowest values (sensitivity 0.414, specificity 0.674).

The model ranked participants well by MCI risk: the proportion with MCI increased from 6% in the lowest quintile of predicted risk to 75% in the highest (Figure 4C). It overestimated risk for all 3 outcomes, most for frailty (mean predicted probability 0.422 vs prevalence 0.278). The Brier score was lower than the reference only for MCI (0.203 vs 0.247), not for frailty or fall risk. All 3 outcomes were predicted correctly for 32.9% of participants (subset accuracy; Multimedia Appendix 4, Table S8 and S9 and Figures S2 and S3).

### Model Interpretation

To understand how the unified model predicted all 3 conditions, we examined which features contributed most and whether they were shared across conditions.

The explanations were consistent between training and unseen test participants: the same 10 features ranked highest in both, 19 of the 20 most important features were the same, and the rankings of all 59 features were almost identical (Spearman ρ=0.99, P<.001; Multimedia Appendix 5, Figure S4 and Table S10). Figure 5 therefore shows the test participants.

**Figure 5.**
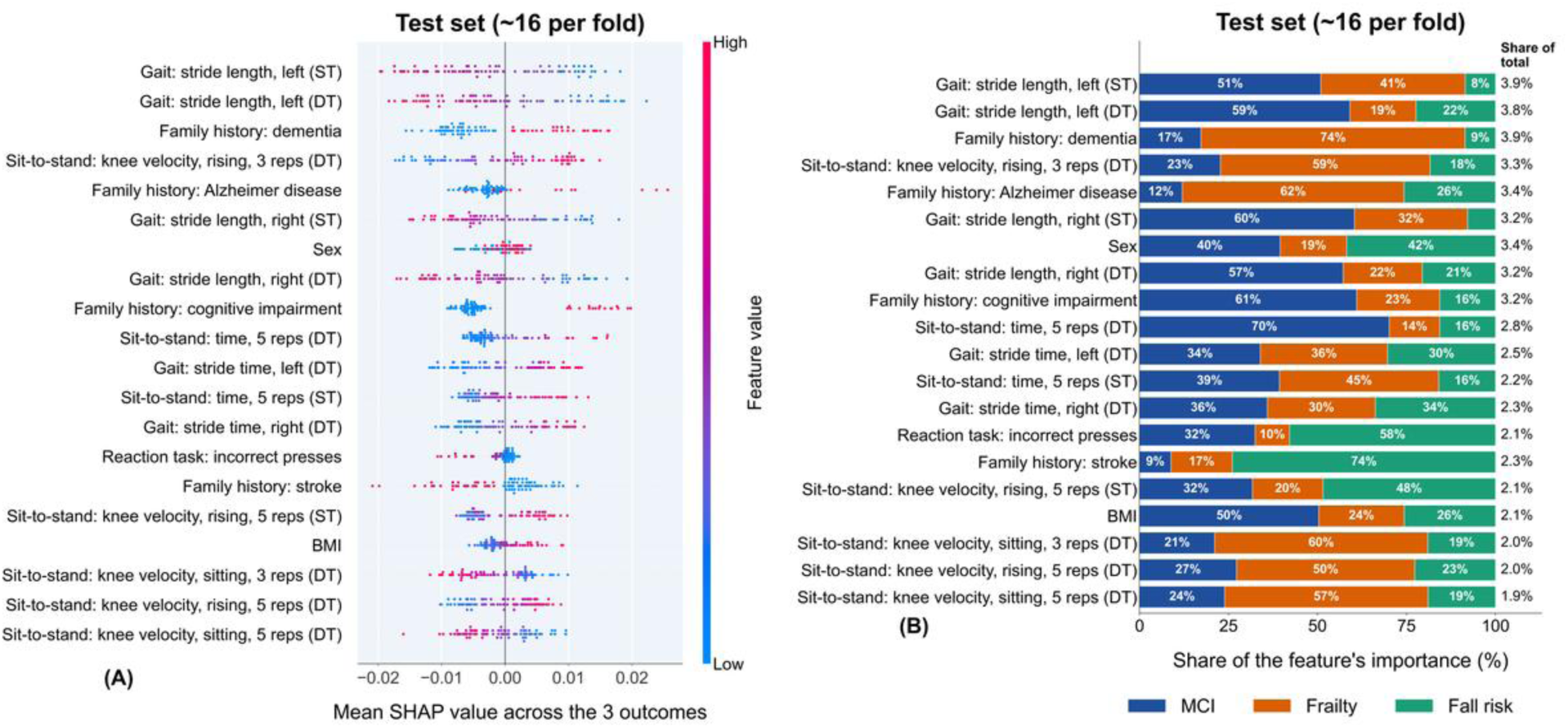
Most important features of the final unified model and their share across MCI, frailty, and fall risk in the test participants. (A) SHAP beeswarm summary plot of the 20 most important features, ranked by mean absolute SHAP value, with SHAP values averaged over the 3 outcomes. Each dot represents a single participant; dots to the right of 0 indicate a higher predicted risk. Dot color shows the feature value, from blue (low) to pink (high); for binary features, pink indicates female or yes. (B) Share of each outcome in each feature’s importance: blue, MCI; orange, frailty; green, fall risk. The feature’s share of total importance is shown at the right.

Gait and STS measures contributed most to the predictions. STS features accounted for 24.3% of the model’s total importance and gait features for 21.8%, followed by family history (16.7%) and static balance (12.8%; training data, with the same order in the test participants). Stride length was the most important single measure (14.2% of total importance), and 9 of the 20 most important features were measured under dual-task conditions (Figure 5A). Averaged across the 3 outcomes, shorter stride length, longer stride time, longer STS time, and a family history of dementia, cognitive impairment, or Alzheimer disease were linked to a higher predicted risk. A family history of stroke was linked to a lower predicted risk. Sex ranked seventh and had opposite effects: male sex was linked to a higher predicted risk of MCI, and female sex to a higher predicted risk of frailty and fall risk.

Most features were shared across the 3 conditions. Of the 59 features, 43 (73%) were shared and accounted for 65.3% of the model’s total importance in the training participants; the other 16 were mainly linked to a single outcome (Multimedia Appendix 5, Table S10). Among the 20 most important features, 13 were shared, including sex and the 2 most important features, left stride length in single-task and dual-task walking (Figure 5B). The 16 features mainly linked to a single outcome included right stride length in single-task walking, dual-task STS time, reaction time, and a family history of cognitive impairment for MCI; dual-task STS knee velocity while sitting and a family history of dementia or Alzheimer disease for frailty; and a family history of stroke or alcoholism for fall risk (Multimedia Appendix 5, Table S10).

### Effect Sizes Between Participants With and Without Each Condition

Group differences followed the same order as the AUCs of the final model (0.779, 0.606, and 0.551): across all 59 features, the median absolute SMD was largest for MCI (0.383), smaller for frailty (0.349), and smallest for fall risk (0.156) (Figure 6A; Multimedia Appendix 5, Tables S11 and S12).

**Figure 6.**
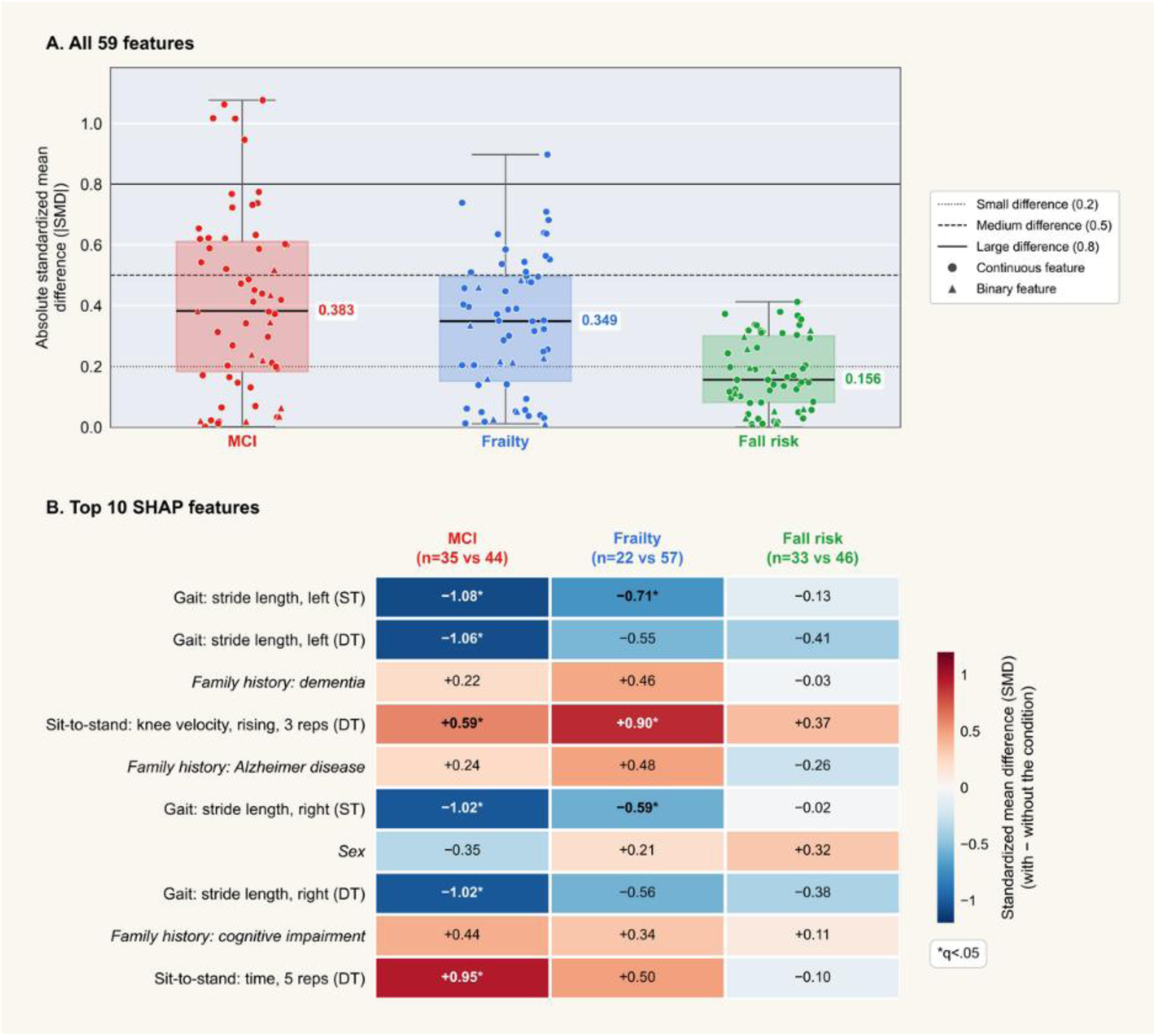
Effect sizes of the features between participants with and without MCI, frailty, and fall risk. (A) Absolute SMDs of all 59 features for MCI (red), frailty (blue), and fall risk (green). Each point is 1 feature (circles, continuous; triangles, binary); boxes show the median and IQR. Dotted, dashed, and solid lines mark small (0.2), medium (0.5), and large (0.8) differences. (B) SMDs, with direction, of the 10 most important features in Figure 5. Red indicates higher values, and blue lower values, in participants with the condition; binary features are in italics. *q<.05. IQR: interquartile range.

The 10 most important features in Figure 5 showed the same pattern (Figure 6B). For MCI, 6 of the 10 differed significantly: participants with MCI had shorter stride lengths, with large differences (SMD −1.02 to −1.08), higher dual-task STS knee velocity while rising (SMD 0.59), and longer dual-task STS time (SMD 0.95). For frailty, 3 differed significantly: higher dual-task STS knee velocity while rising (SMD 0.90) and shorter single-task stride lengths (SMD −0.71 and −0.59). For fall risk, no feature differed significantly, and all differences were small (absolute SMD ≤0.41). These differences were in the same direction as the SHAP effects; for example, shorter stride length was linked both to MCI and to a higher predicted risk. The family-history features contributed to the predictions without significant group differences on their own.

### Robustness Analyses

If the 3 conditions are linked, the chain should use the prediction for one condition to inform the next. We tested whether it did, and whether performance depended on any single feature domain or on the order of the chain.

#### Chain-Input Importance

The chain used the link between frailty and fall risk. When predicted frailty was shuffled among the test participants, the fall-risk AUC decreased by 0.062 (SD 0.013), making predicted frailty the most important of the 61 inputs of the fall-risk step, with an effect 3.5 times that of the most important original feature (reaction-task incorrect presses, 0.017) (Figure 7A; Multimedia Appendix 5, Table S13). Predicted MCI contributed less: it ranked 17th of 61 inputs for fall risk (0.004) and 5th of 60 for frailty, where its effect (0.006) was about half that of the most important original feature (0.011).

**Figure 7.**
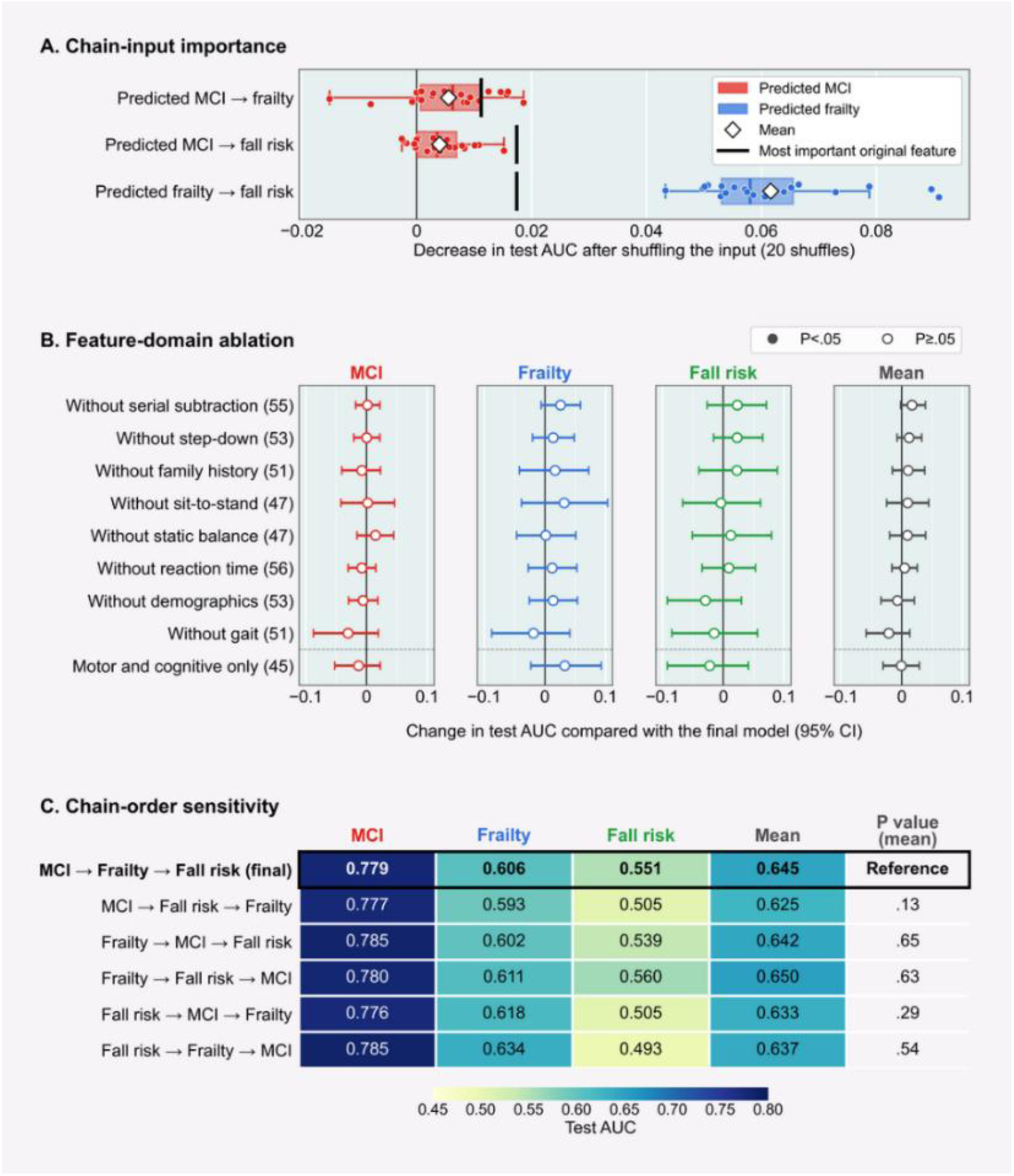
Robustness of the final unified model to chain inputs, feature domains, and chain order. (A) Decrease in test AUC when each chain prediction was shuffled (20 shuffles; black line, most important original feature). (B) Change in test AUC (95% CI) after removing each domain (features kept in parentheses). (C) Test AUC for the 6 chain orders; P values compare the mean AUC of each order with the final order (outlined).

#### Feature-Domain Ablation

No single feature domain was essential (Figure 7B): removing any 1 domain changed the mean test AUC by −0.022 to 0.017, and no change was significant. A model using only the 45 sensor features from the motor and cognitive tasks performed as well as the final model (mean AUC 0.645; change −0.001, 95% CI −0.031 to 0.029). Removing gait features caused the largest decrease (0.624; change −0.022, 95% CI −0.058 to 0.013) and serial-subtraction features the largest increase (0.662; change 0.017, 95% CI −0.002 to 0.039) (Multimedia Appendix 5, Table S14).

#### Chain-Order Sensitivity

The order of the outcomes in the chain did not significantly change overall performance (Figure 7C): mean test AUC ranged from 0.625 to 0.650 across the 6 orders (P=.13 to.65 vs the final order), and MCI AUC was consistent (0.776-0.785). Fall-risk AUC, however, depended on where frailty sat in the chain: 0.539-0.560 when frailty preceded fall risk but 0.493-0.505 when it followed, with the only significant difference for the order fall risk → frailty → MCI (0.493 vs 0.551; P=.03) (Multimedia Appendix 5, Table S15). Fall-risk prediction therefore benefited from predicting frailty first.

### Sample-Size Analyses

Frailty and fall risk remained harder to predict than MCI, so we examined whether the sample was large enough. Learning curves showed whether performance would improve with more participants, and a sensitivity power analysis showed how large a group difference the sample could reliably detect.

#### Learning Curve

Mean test AUC increased from 0.615 with 25 training participants to 0.639 with 44 and then changed little (0.640 with 53 and 0.645 with 63; Figure 8A); no smaller training size differed significantly from the full size (all P≥.12). The outcomes, however, followed different patterns. MCI AUC increased from 0.736 to 0.770 at 44 participants and then leveled off (0.779 at 63), and frailty AUC peaked at 0.623 at 44 participants and then decreased slightly (0.606 at 63). Fall-risk AUC remained near chance (0.524-0.533) up to 53 participants and reached 0.551 only at 63, making fall risk the only outcome still improving at the largest training size. Training AUC remained close to 1 at every size (0.978-0.993), well above the test AUC, indicating that the model fitted the training participants much more closely than new participants.

**Figure 8.**
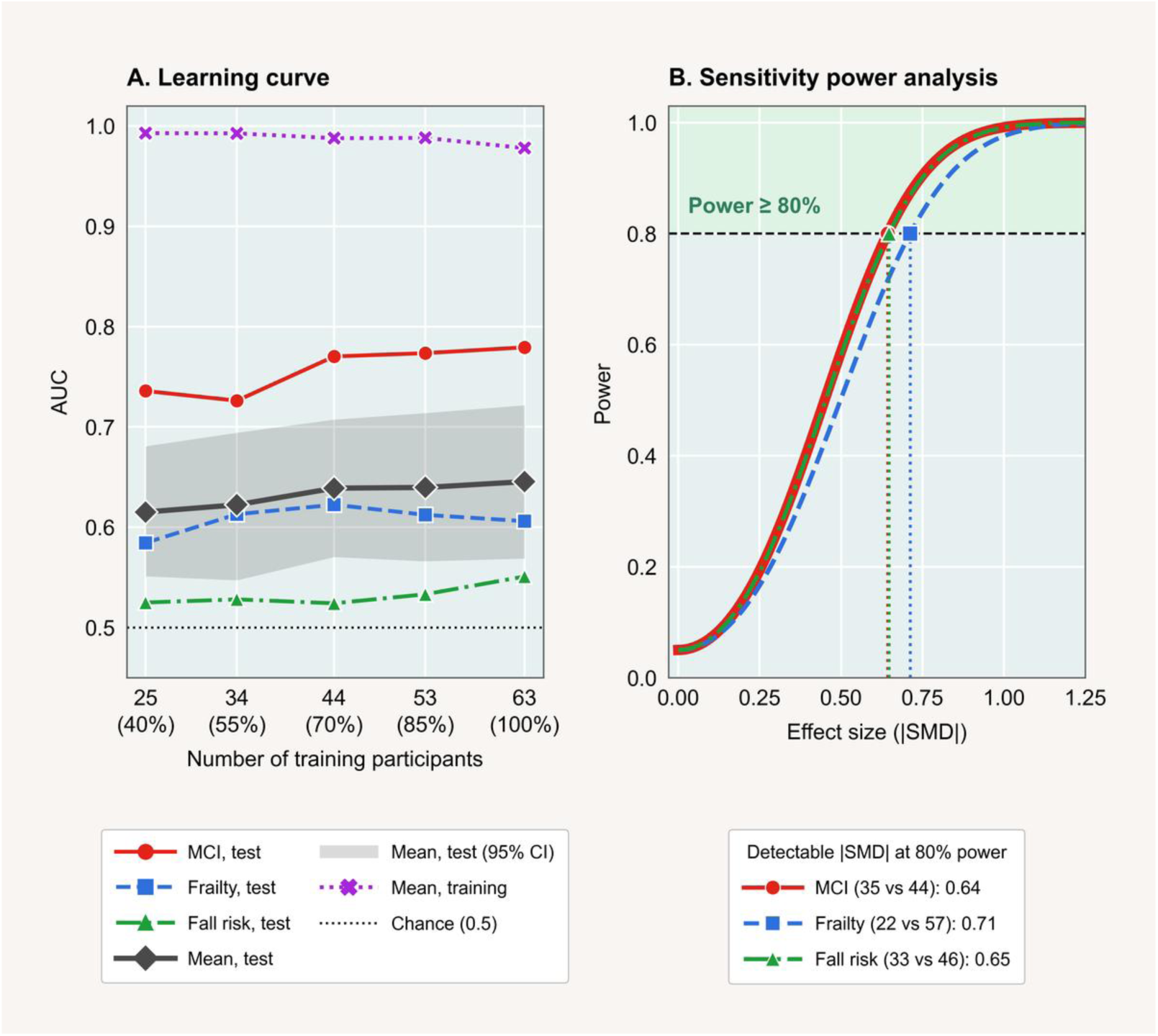
Learning curve and sensitivity power analysis of the final unified model. (A) Test AUC for each outcome and the mean (grey band, 95% CI), and mean training AUC, by number of training participants. (B) Power to detect an absolute SMD (2-sided t test, α=.05); dotted lines mark the smallest SMD detectable with 80% power.

#### Sensitivity Power Analysis

With 79 participants, the smallest group difference detectable with 80% power was an absolute SMD of 0.64 for MCI, 0.65 for fall risk, and 0.71 for frailty (Figure 8B; Multimedia Appendix 6, Table S16), so the study could reliably detect only medium-to-large effects. The power to detect a medium effect (SMD 0.5) was 0.50-0.59, and detecting such an effect with 80% power would require about 130 participants for MCI, 132 for fall risk, and 159 for frailty.

## Discussion

### Principal Findings

This study tested whether a unified model could screen older adults for MCI, frailty, and fall risk at once from one motor-function assessment. The findings support 3 conclusions.

First, the unified classifier-chain model performed as well as 3 separate single-label models. With class weighting, it achieved a higher mean AUC in 17 of 20 combinations of algorithm and stage, both significant differences showed a higher AUC for the unified model, and it performed at least as well under nested model selection. It was also more robust to feature selection than the single-label models. ET with all features gave the best unified model, consistent with its random split thresholds, which reduce variance in small samples [40]. The final model discriminated MCI well (AUC 0.779) and ranked participants by MCI risk: the proportion with MCI increased from 6% in the lowest quintile of predicted risk to 75% in the highest. Frailty (AUC 0.606) and fall risk (0.551) were discriminated less well, and risk was overestimated for all 3 outcomes. Three findings indicate that the weaker results for frailty and fall risk reflect the data rather than the unified approach: single-label models were equally weak for these outcomes, the group differences in the features were smallest for these outcomes, and fall-risk AUC was still increasing at the largest training size.

Second, the explanations were consistent and clinically meaningful. The same features ranked highest in training participants and in unseen test participants, and gait and STS features contributed most, led by stride length, which also showed large group differences for MCI (SMD −1.02 to −1.08). Most features (73%), including the 2 most important, were shared across the 3 conditions and provided two-thirds of the model’s importance. This shared motor basis is what allows a unified model to predict all 3 conditions.

Third, the chain used the link between the conditions. Predicted frailty was the most important of the 61 inputs for fall risk, with an effect 3.5 times that of any original feature, and fall-risk AUC was lowest when fall risk was predicted before frailty. Overall performance did not depend on the chain order or on any single feature domain, and the 45 sensor features alone performed as well as the full model.

### Comparison With Previous Work

Few studies have predicted several health outcomes with a unified model. Zhou et al [25] predicted 4 diabetic complications from electronic health records of 9765 patients (AUC 0.823), and Tarekegn et al [26] predicted 6 frailty-related outcomes from the administrative data of 105,962 people, with a higher macro-AUC for the multi-label model than for separate models (0.83 vs 0.78). Our results are consistent with these studies: the unified model had a higher AUC in most configurations and never a significantly lower one when class imbalance was handled with class weighting. We extend this approach to MCI, frailty, and fall risk and to sensor-based motor-function data. Our sample was more than 100 times smaller and used a single assessment rather than broad clinical records, which may partly explain the lower AUCs.

The motor features in our model have mostly been studied for one condition at a time: stride length and double support separated frailty levels [56], wearable motion sensors provided indicators of fall risk [57], and dual-task gait has been associated with progression from MCI to dementia [16], and with future falls [58]. Our finding that most features were shared across the 3 conditions is consistent with their clinical overlap. Motoric cognitive risk syndrome, defined by slow gait and cognitive complaints, was present in nearly 10% of more than 26,000 older adults and predicted dementia [59], and cognitive frailty describes the co-occurrence of physical frailty and cognitive impairment [60]. Frailty is also an established predictor of falls [13]. Classifier chains are designed to use such dependencies between outcomes [24], and our chain analyses show that the model used this dependency: predicted frailty was the most important input for fall risk.

Two findings show why a unified model needs outcome-specific interpretation. First, a feature can affect the outcomes in opposite directions, which averaging across outcomes would hide: male sex increased the predicted risk of MCI, whereas female sex increased that of frailty and fall risk. Second, family-history features and sex ranked high in SHAP but showed no significant group differences, and removing demographics and family history did not change performance (mean AUC 0.645). These features therefore did not add predictive information beyond the motor features, and their SHAP importance should not be interpreted as evidence of risk.

Our class-imbalance results are consistent with those of van den Goorbergh et al [47], who showed that imbalance corrections, including SMOTE, did not improve discrimination and reduced calibration. In our study, SMOTE reduced the AUC of the unified model in all 10 combinations, and even class weighting led to overestimated risks, most for frailty.

## Strengths and Limitations

To our knowledge, this is the first study to compare a unified model for MCI, frailty, and fall risk with single-label models under the identical settings. It quantified each feature’s importance for each outcome, tested the explanations on unseen participants, linked them to group differences, and showed through permutation analysis that the chain used the dependency between frailty and fall risk. Repeated nested cross-validation with bootstrap CIs ensured that performance was always estimated on unseen participants and reported with its uncertainty.

The study also has limitations. The sample was small (n=79), recruited at a single institution, and almost all White, which limits precision and generalizability; external validation in independent cohorts is needed. Fall risk was based on self-reported past falls, so the model identifies fall history rather than future falls, and self-reported falls and family history are subject to recall error. MCI was partly defined by the MoCA, which includes serial subtraction, and the frailty questionnaire includes memory and balance items; these overlaps can link some outcomes and features. Discrimination was modest for frailty and close to chance for fall risk, and the model overestimated risk, so it requires improvement and recalibration before clinical use. Because the study was cross-sectional, the model identifies current conditions rather than predicting their onset. Finally, SHAP values describe how the model uses the features, not causal effects, and the 60% threshold for features linked mainly to a single outcome was our own choice.

### Clinical Implications and Future Directions

One motor-function assessment could support screening for MCI, frailty, and fall risk at the same visit. The 45 sensor features alone performed as well as the full model, so screening could rely on the MPASS recording alone, without demographic or family-history information. Because MPASS is portable and its depth-camera measurements of gait and STS have been validated [35,36], this screening could move from specialist clinics to primary care and community settings. For health informatics, a unified model replaces 3 separate models to develop, validate, deploy, and update, and its outcome-specific SHAP explanations show how each feature contributes to each prediction. In its current form, the model is most useful for MCI. Future studies should validate and recalibrate it in a larger and more diverse cohort, follow participants over time to test whether it predicts the onset of these conditions, and measure whether such screening reduces assessment time and costs. Fall-risk AUC was still increasing at the largest training size, so fall-risk prediction has the most to gain from a larger sample.

## Conclusions

A unified classifier-chain model predicted MCI, frailty, and fall risk together from motor-function data and performed as well as 3 separate models when class imbalance was handled with class weighting. It identified MCI well, but frailty and fall risk less reliably. Its explanations were consistent and showed that most motor features were shared across the 3 conditions, and the chain used the link between frailty and fall risk. With external validation in larger cohorts, such a model could support efficient screening of older adults for several conditions from a single motor-function assessment.

## Supporting information

Multimedia Appendices 1-5

## Data Availability

The data analyzed in this study are not publicly available because they contain information from human participants, but de-identified data are available from the corresponding author upon reasonable request and with approval from the University of Missouri Institutional Review Board.

## Acknowledgements

We thank the older adults who participated in this study and the staff of the Mizzou Motion Analysis Center for their help with data collection.

## Authors’ Contributions

Conceptualization: SA, TMG, PR

Data curation: SA, TMG, SS, SAH, JBH

Formal analysis: SA

Funding acquisition: TMG, AMK, JBH, PR

Methodology: SA, PR

Supervision: PR, TMG

Visualization: SA

Writing – original draft: SA

Writing – review & editing: SA, TMG, PR

## Initials

SA = Sonia Akter, TMG = Trent M. Guess, SS = Shraboni Sarker, SAH = Samuel A. Hockett, AMK = Andrew M. Kiselica, JBH = Jamie B. Hall, PR = Praveen Rao.

## Data Availability

The datasets analyzed during this study are not publicly available because they contain information that could compromise participant privacy, but they are available from the corresponding author upon reasonable request. The analysis code is available from the first author (SA) upon reasonable request.

## Abbreviations

AP: anteroposterior
AUC: area under the receiver operating characteristic curve
BMI: body mass index
CoM: center of mass
CoP: center of pressure
DT: dual task
ET: extra trees
GB: gradient boosting
LR: logistic regression
MCI: mild cognitive impairment
ML (in features): mediolateral
ML: machine learning
MoCA: Montreal Cognitive Assessment
MPASS: Mizzou Point-of-Care Assessment System
NPV: negative predictive value
PPV: positive predictive value
PR-AUC: area under the precision-recall curve
RF: random forest
RFE: recursive feature elimination
ROC: receiver operating characteristic
SHAP: Shapley additive explanations
SMD: standardized mean difference
SMOTE: synthetic minority oversampling technique
ST: single task
STS: sit-to-stand
XGBoost: extreme gradient boosting

