## Supplementary material for "Unified Machine Learning Model for Early Detection of Mild Cognitive Impairment, Frailty, and Fall Risk Using Multimodal Motor-Function Data": Multimedia Appendices 1-5

#### Multimedia Appendix. Features used in the models

**Table S1.** The 59 features used in the models, by domain

| Domain | Feature (as named in figures) | Description | Condition | Unit | Source |
|---|---|---|---|---|---|
| **Demographics** | Age | Age at assessment | — | years | Questionnaire |
| | Height | Body height | — | m | Questionnaire |
| | Weight | Body weight | — | kg | Questionnaire |
| | BMI | Body mass index | — | $kg/m^2$ | Calculated |
| | Sex | 0=male, 1=female | — | — | Questionnaire |
| | Race | 0=White, 1=non-White | — | — | Questionnaire |
| **Family history** | Family history: Alzheimer disease | Self-reported family history of Alzheimer disease; 0=no, 1=yes | — | — | Questionnaire |
| | Family history: dementia | Self-reported family history of dementia; 0=no, 1=yes | — | — | Questionnaire |
| | Family history: Parkinson disease | Self-reported family history of Parkinson disease; 0=no, 1=yes | — | — | Questionnaire |
| | Family history: stroke | Self-reported family history of stroke; 0=no, 1=yes | — | — | Questionnaire |
| | Family history: cognitive impairment | Self-reported family history of cognitive impairment; 0=no, 1=yes | — | — | Questionnaire |
| | Family history: schizophrenia | Self-reported family history of schizophrenia; 0=no, 1=yes | — | — | Questionnaire |
| | Family history: bipolar disorder | Self-reported family history of bipolar disorder; 0=no, 1=yes | — | — | Questionnaire |
| | Family history: alcoholism | Self-reported family history of alcoholism; 0=no, 1=yes | — | — | Questionnaire |
| **Gait** | Gait: stride length, left (ST) | Mean stride length of the left foot | Single task | m | Depth camera |
| | Gait: stride length, right (ST) | Mean stride length of the right foot | Single task | m | Depth camera |
| | Gait: stride time, left (ST) | Mean stride time of the left foot | Single task | s | Depth camera |
| | Gait: stride time, right (ST) | Mean stride time of the right foot | Single task | s | Depth camera |
| | Gait: stride length, left (DT) | Mean stride length of the left foot | Dual task | m | Depth camera |
| | Gait: stride length, right (DT) | Mean stride length of the right foot | Dual task | m | Depth camera |
| | Gait: stride time, left (DT) | Mean stride time of the left foot | Dual task | s | Depth camera |
| | Gait: stride time, right (DT) | Mean stride time of the right foot | Dual task | s | Depth camera |
| **Sit-to-stand** | Sit-to-stand: time, 3 reps (ST) | Time to complete 3 sit-to-stand repetitions | Single task | s | Depth camera |
| | Sit-to-stand: knee velocity, rising, 3 reps (ST) | Mean knee angular velocity while rising, over 3 repetitions | Single task | deg/s | Depth camera |
| | Sit-to-stand: knee velocity, sitting, 3 reps (ST) | Mean knee angular velocity while sitting down, over 3 repetitions | Single task | deg/s | Depth camera |

| Domain | Feature (as named in figures) | Description | Condition | Unit | Source |
| --- | --- | --- | --- | --- | --- |
| | Sit-to-stand: time, 5 reps (ST) | Time to complete 5 sit-to-stand repetitions | Single task | s | Depth camera |
| | Sit-to-stand: knee velocity, rising, 5 reps (ST) | Mean knee angular velocity while rising, over 5 repetitions | Single task | deg/s | Depth camera |
| | Sit-to-stand: knee velocity, sitting, 5 reps (ST) | Mean knee angular velocity while sitting down, over 5 repetitions | Single task | deg/s | Depth camera |
| | Sit-to-stand: time, 3 reps (DT) | Time to complete 3 sit-to-stand repetitions | Dual task | s | Depth camera |
| | Sit-to-stand: knee velocity, rising, 3 reps (DT) | Mean knee angular velocity while rising, over 3 repetitions | Dual task | deg/s | Depth camera |
| | Sit-to-stand: knee velocity, sitting, 3 reps (DT) | Mean knee angular velocity while sitting down, over 3 repetitions | Dual task | deg/s | Depth camera |
| | Sit-to-stand: time, 5 reps (DT) | Time to complete 5 sit-to-stand repetitions | Dual task | s | Depth camera |
| | Sit-to-stand: knee velocity, rising, 5 reps (DT) | Mean knee angular velocity while rising, over 5 repetitions | Dual task | deg/s | Depth camera |
| | Sit-to-stand: knee velocity, sitting, 5 reps (DT) | Mean knee angular velocity while sitting down, over 5 repetitions | Dual task | deg/s | Depth camera |
| **Static balance** | Balance: CoP 95% sway area (ST) | Area of the 95% prediction ellipse of CoP sway | Single task | $mm^2$ | Force plate |
| | Balance: CoP mean velocity (ST) | Mean CoP velocity (total) | Single task | mm/s | Force plate |
| | Balance: CoP mean velocity, ML (ST) | Mean CoP velocity in the mediolateral direction | Single task | mm/s | Force plate |
| | Balance: CoP mean velocity, AP (ST) | Mean CoP velocity in the anteroposterior direction | Single task | mm/s | Force plate |
| | Balance: AUC, AP (ST) | Area-under-the-curve measure of anteroposterior CoP displacement | Single task | — | Force plate |
| | Balance: AUC, ML (ST) | Area-under-the-curve measure of mediolateral CoP displacement | Single task | — | Force plate |
| | Balance: CoP 95% sway area (DT) | Area of the 95% prediction ellipse of CoP sway | Dual task | $mm^2$ | Force plate |
| | Balance: CoP mean velocity (DT) | Mean CoP velocity (total) | Dual task | mm/s | Force plate |
| | Balance: CoP mean velocity, ML (DT) | Mean CoP velocity in the mediolateral direction | Dual task | mm/s | Force plate |
| | Balance: CoP mean velocity, AP (DT) | Mean CoP velocity in the anteroposterior direction | Dual task | mm/s | Force plate |
| | Balance: AUC, AP (DT) | Area-under-the-curve measure of anteroposterior CoP displacement | Dual task | — | Force plate |
| | Balance: AUC, ML (DT) | Area-under-the-curve measure of mediolateral CoP displacement | Dual task | — | Force plate |
| **Step-down** | Step-down: lead time, left | Step-down time of the leading limb, left step-down | — | s | Depth camera |
| | Step-down: trail time, left | Step-down time of the trailing limb, left step-down | — | s | Depth camera |
| | Step-down: CoM ML displacement, left | Mediolateral center-of-mass displacement, left step-down | — | mm | Depth camera |
| | Step-down: lead time, right | Step-down time of the leading limb, right step-down | — | s | Depth camera |

| Domain | Feature (as named in figures) | Description | Condition | Unit | Source |
|---|---|---|---|---|---|
| | Step-down: trail time, right | Step-down time of the trailing limb, right step-down | — | s | Depth camera |
| | Step-down: CoM ML displacement, right | Mediolateral center-of-mass displacement, right step-down | — | mm | Depth camera |
| **Reaction time** | Reaction task: reaction time | Mean time to respond to light cues | — | ms | Interface board |
| | Reaction task: incorrect presses | Number of incorrect button presses | — | count | Interface board |
| | Reaction task: buttons pressed | Total number of button presses | — | count | Interface board |
| **Serial subtraction** | Serial subtraction: total responses, 15 s (DT) | Number of responses in 15 s during dual-task standing | Dual task | count | Static-balance DT trials |
| | Serial subtraction: incorrect responses, 15 s (DT) | Number of incorrect responses in 15 s during dual-task standing | Dual task | count | Static-balance DT trials |
| | Serial subtraction: interference, total (DT) | Dual-task interference on total responses (percentage change from single task) | Dual task | % | Static-balance DT trials |
| | Serial subtraction: interference, correct (DT) | Dual-task interference on correct responses (percentage change from single task) | Dual task | % | Static-balance DT trials |

Sensor features were averaged over repeated trials. CoP: center of pressure; CoM: center of mass; AP: anteroposterior; ML: mediolateral; ST: single task; DT: dual task (serial subtraction). Feature names match those in Figures 5 and 6.

**Multimedia Appendix. Hyperparameter search spaces**

**Table S2. Hyperparameter search spaces for the unified and single-label models**

| Algorithm | Hyperparameter evaluated |
|---|---|
| LR | C = {0.01, 0.1, 1, 10, 100}, Penalty = {L1, L2} |
| RF | n_estimators = {200, 300}, max_depth = {3, 5, 7, None}, min_samples_split = {2, 5}, min_samples_leaf = {1, 2} |
| ET | n_estimators = {100, 200, 300}, max_depth = {3, 5, 7, None}, min_samples_split = {2, 5, 10}, min_samples_leaf = {1, 2, 3}, max_features = {sqrt, log2}, bootstrap = {True, False}, criterion = {gini, entropy} |
| GB | n_estimators = {100, 200, 300}, learning_rate = {0.01, 0.03, 0.05}, max_depth = {1, 2, 3}, min_samples_leaf = {1, 2, 3}, subsample = {0.8, 1.0} |
| XGBoost | n_estimators = {100, 200, 300}, learning_rate = {0.01, 0.03, 0.05}, max_depth = {1, 2, 3}, min_child_weight = {1, 3}, subsample = {0.8, 1.0}, colsample_bytree = {0.8, 1.0}, gamma = {0, 0.1}, reg_lambda = {1, 5} |

ET: extra trees; GB: gradient boosting; LR: logistic regression; RF: random forest; XGBoost: extreme gradient boosting. When a search space contained 50 or fewer combinations, all combinations were evaluated; otherwise, 50 combinations were sampled at random.

### Multimedia Appendix. Correlation between the features

#### Figure S1. Pearson correlation matrix of the 59 features, grouped by domain

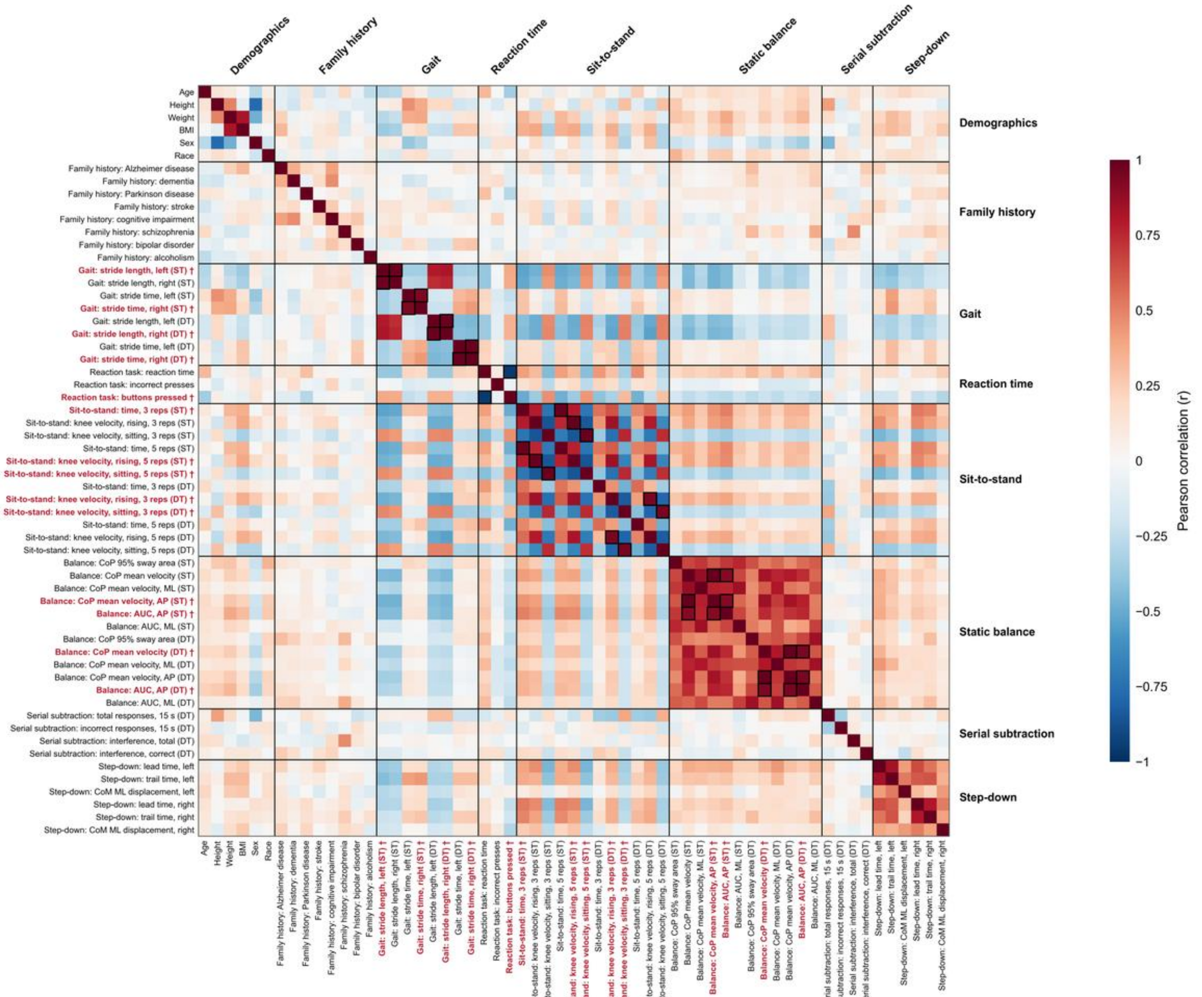


Black boxes mark feature pairs with |r|>0.90, and red labels mark the 14 features removed by the correlation filter (stage S2) when it was applied to all 79 participants. Because the filter was refitted within each training set, the features removed could differ slightly between cross-validation splits. AP: anteroposterior; CoM: center of mass; CoP: center of pressure; DT: dual task; ML: mediolateral; reps: repetitions; ST: single task.

#### Multimedia Appendix. Participant characteristics and supplementary performance results

#### Table S3. Characteristics of the participants by outcome group (N=79)

| MCI<br>Characteristic | Overall (n=79) | Without MCI (n=44) | With MCI (n=35) | P value |
| --- | --- | --- | --- | --- |
| Age (years), mean (SD) | 70.86 (4.68) | 69.93 (4.08) | 72.03 (5.17) | .054 |
| Height (m), mean (SD) | 1.68 (0.10) | 1.69 (0.09) | 1.68 (0.11) | .78 |
| Weight (kg), mean (SD) | 79.48 (16.60) | 75.57 (13.88) | 84.40 (18.53) | .02[a] |
| BMI (kg/m²), mean (SD) | 27.98 (5.21) | 26.53 (4.58) | 29.81 (5.43) | .006[a] |
| Sex, n (%) | | | | .17 |
| Female | 48 (60.8) | 30 (68.2) | 18 (51.4) | |
| Male | 31 (39.2) | 14 (31.8) | 17 (48.6) | |
| Race, n (%) | | | | >.99 |
| White | 77 (97.5) | 43 (97.7) | 34 (97.1) | |
| Black/African American | 2 (2.5) | 1 (2.3) | 1 (2.9) | |
| **Frailty**<br>**Characteristic** | **Overall (n=79)** | **Without frailty (n=57)** | **With frailty (n=22)** | **P value** |
| Age (years), mean (SD) | 70.86 (4.68) | 70.88 (4.08) | 70.82 (6.08) | .97 |
| Height (m), mean (SD) | 1.68 (0.10) | 1.69 (0.09) | 1.66 (0.12) | .21 |
| Weight (kg), mean (SD) | 79.48 (16.60) | 78.83 (16.05) | 81.17 (18.23) | .60 |
| BMI (kg/m²), mean (SD) | 27.98 (5.21) | 27.41 (5.08) | 29.47 (5.36) | .13 |
| Sex, n (%) | | | | .45 |
| Female | 48 (60.8) | 33 (57.9) | 15 (68.2) | |
| Male | 31 (39.2) | 24 (42.1) | 7 (31.8) | |
| Race, n (%) | | | | .48 |
| White | 77 (97.5) | 56 (98.2) | 21 (95.5) | |
| Black/African American | 2 (2.5) | 1 (1.8) | 1 (4.5) | |
| **Fall risk**<br>**Characteristic** | **Overall (n=79)** | **Without fall risk (n=46)** | **With fall risk (n=33)** | **P value** |
| Age (years), mean (SD) | 70.86 (4.68) | 71.52 (5.09) | 69.94 (3.93) | .12 |
| Height (m), mean (SD) | 1.68 (0.10) | 1.69 (0.10) | 1.67 (0.10) | .38 |
| Weight (kg), mean (SD) | 79.48 (16.60) | 81.83 (15.66) | 76.21 (17.54) | .15 |
| BMI (kg/m²), mean (SD) | 27.98 (5.21) | 28.67 (5.55) | 27.03 (4.60) | .16 |
| Sex, n (%) | | | | .24 |
| Female | 48 (60.8) | 25 (54.3) | 23 (69.7) | |
| Male | 31 (39.2) | 21 (45.7) | 10 (30.3) | |
| Race, n (%) | | | | >.99 |
| White | 77 (97.5) | 45 (97.8) | 32 (97.0) | |
| Black/African American | 2 (2.5) | 1 (2.2) | 1 (3.0) | |

[a]$P<.05$.

P values were calculated with one-way ANOVA for continuous variables and the chi-square or Fisher exact test for categorical variables.

BMI: body mass index; MCI: mild cognitive impairment.

**Table S4. Mean AUC of the unified and single-label approaches for every algorithm and stage (S1–S6)**

| Stage | Algorithm | Unified | Single-label | Difference (95% CI) | *P* |
|---|---|---|---|---|---|
| S1 | LR | 0.604 | 0.565 | +0.040 (0.000 to +0.079) | .052 |
| | RF | 0.643 | 0.648 | −0.004 (−0.019 to +0.012) | .56 |
| | ET | 0.645 | 0.625 | +0.020 (−0.009 to +0.045) | .15 |
| | XGB | 0.645 | 0.628 | +0.018 (−0.003 to +0.037) | .09 |
| | GB | 0.635 | 0.627 | +0.008 (−0.023 to +0.040) | .62 |
| S2 | LR | 0.572 | 0.555 | +0.017 (−0.030 to +0.058) | .43 |
| | RF | 0.636 | 0.621 | +0.014 (−0.002 to +0.033) | .08 |
| | ET | 0.626 | 0.606 | +0.020 (−0.006 to +0.047) | .14 |
| | XGB | 0.633 | 0.623 | +0.010 (−0.009 to +0.030) | .26 |
| | GB | 0.630 | 0.617 | +0.013 (−0.019 to +0.045) | .43 |
| S3 | LR | 0.579 | 0.572 | +0.007 (−0.034 to +0.053) | .73 |
| | RF | 0.634 | 0.620 | +0.014 (−0.011 to +0.040) | .29 |
| | ET | 0.637 | 0.620 | +0.017 (−0.017 to +0.050) | .31 |
| | XGB | 0.639 | 0.608 | **+0.031 (+0.001 to +0.061)** | **.048** |
| | GB | 0.603 | 0.604 | −0.000 (−0.035 to +0.033) | .90 |
| S4 | LR | 0.619 | 0.588 | +0.031 (−0.019 to +0.082) | .21 |
| | RF | 0.620 | 0.603 | +0.017 (−0.015 to +0.050) | .33 |
| | ET | 0.633 | 0.583 | **+0.050 (+0.013 to +0.089)** | **.01** |
| | XGB | 0.627 | 0.607 | +0.020 (−0.008 to +0.050) | .19 |
| | GB | 0.586 | 0.595 | −0.009 (−0.041 to +0.021) | .54 |
| S5 | LR | 0.565 | 0.577 | −0.012 (−0.043 to +0.024) | .52 |
| | RF | 0.589 | 0.625 | −0.036 (−0.075 to +0.002) | .07 |
| | ET | 0.581 | 0.624 | **−0.042 (−0.086 to −0.005)** | **.03** |
| | XGB | 0.570 | 0.622 | **−0.051 (−0.091 to −0.016)** | **.008** |
| | GB | 0.557 | 0.624 | **−0.067 (−0.122 to −0.015)** | **.01** |
| S6 | LR | 0.526 | 0.577 | **−0.051 (−0.096 to −0.003)** | **.03** |
| | RF | 0.559 | 0.605 | −0.046 (−0.093 to 0.000) | .052 |
| | ET | 0.553 | 0.583 | −0.030 (−0.068 to +0.006) | .12 |
| | XGB | 0.543 | 0.589 | −0.046 (−0.102 to +0.008) | .09 |
| | GB | 0.545 | 0.583 | −0.038 (−0.093 to +0.008) | .14 |

Mean AUC across MCI, frailty, and fall risk; pooled out-of-fold predictions of the nested cross-validation (79 participants, 3 repeats). Difference: unified − single-label; 95% CIs and P values from the paired bootstrap (1000 resamples). Bold: P<.05. S1–S4 used class weights and S5–S6 used SMOTE; the stages are defined in Table 2 of the main text. Outcome-specific differences are shown in Figure 3 of the main text.

AUC: area under the receiver operating characteristic curve; ET: extra trees; GB: gradient boosting; LR: logistic regression; MCI: mild cognitive impairment; PR-AUC: area under the precision-recall curve; RF: random forest; SMOTE: synthetic minority oversampling technique; XGB: extreme gradient boosting.

**Table S5. Outcome-specific performance of the unified and single-label approaches using all features (S1)**

| Approach | Algorithm | Outcome | AUC | PR-AUC | Sens | Spec | F1w |
|---|---|---|---|---|---|---|---|
| Unified | LR | MCI | 0.767 | 0.682 | 0.724 | 0.765 | 0.746 |
| | | Frailty | 0.560 | 0.395 | 0.530 | 0.596 | 0.598 |
| | | Fall risk | 0.485 | 0.429 | 0.495 | 0.522 | 0.513 |
| | RF | MCI | 0.781 | 0.711 | 0.619 | 0.788 | 0.711 |
| | | Frailty | 0.605 | 0.420 | 0.288 | 0.854 | 0.675 |
| | | Fall risk | 0.543 | 0.449 | 0.384 | 0.659 | 0.538 |
| | ET | MCI | 0.779 | 0.684 | 0.648 | 0.818 | 0.740 |
| | | Frailty | 0.606 | 0.414 | 0.500 | 0.790 | 0.711 |
| | | Fall risk | 0.551 | 0.490 | 0.414 | 0.674 | 0.560 |
| | XGB | MCI | 0.733 | 0.654 | 0.590 | 0.727 | 0.665 |
| | | Frailty | 0.630 | 0.399 | 0.576 | 0.690 | 0.672 |
| | | Fall risk | 0.572 | 0.470 | 0.586 | 0.587 | 0.588 |
| | GB | MCI | 0.744 | 0.652 | 0.581 | 0.742 | 0.668 |
| | | Frailty | 0.619 | 0.392 | 0.439 | 0.708 | 0.640 |
| | | Fall risk | 0.542 | 0.450 | 0.465 | 0.659 | 0.575 |
| Single-label | LR | MCI | 0.750 | 0.691 | 0.686 | 0.742 | 0.717 |
| | | Frailty | 0.480 | 0.305 | 0.500 | 0.450 | 0.490 |
| | | Fall risk | 0.465 | 0.417 | 0.465 | 0.464 | 0.467 |
| | RF | MCI | 0.780 | 0.707 | 0.619 | 0.773 | 0.703 |
| | | Frailty | 0.622 | 0.424 | 0.288 | 0.877 | 0.688 |
| | | Fall risk | 0.541 | 0.445 | 0.333 | 0.717 | 0.541 |
| | ET | MCI | 0.789 | 0.696 | 0.619 | 0.826 | 0.730 |
| | | Frailty | 0.589 | 0.405 | 0.333 | 0.842 | 0.683 |
| | | Fall risk | 0.497 | 0.424 | 0.354 | 0.674 | 0.530 |
| | XGB | MCI | 0.722 | 0.648 | 0.629 | 0.712 | 0.675 |
| | | Frailty | 0.608 | 0.394 | 0.515 | 0.702 | 0.662 |
| | | Fall risk | 0.552 | 0.468 | 0.515 | 0.594 | 0.563 |
| | GB | MCI | 0.744 | 0.655 | 0.638 | 0.727 | 0.687 |
| | | Frailty | 0.558 | 0.325 | 0.424 | 0.696 | 0.630 |
| | | Fall risk | 0.580 | 0.467 | 0.454 | 0.667 | 0.574 |

Pooled out-of-fold predictions of the nested cross-validation (79 participants, 3 repeats). Sens: sensitivity; Spec: specificity; F1w: weighted F1-score (all at a threshold of 0.5). The prevalence of each outcome (the chance-level PR-AUC) was 0.443 for MCI, 0.278 for frailty, and 0.418 for fall risk.

AUC: area under the receiver operating characteristic curve; ET: extra trees; GB: gradient boosting; LR: logistic regression; MCI: mild cognitive impairment; PR-AUC: area under the precision-recall curve; RF: random forest; SMOTE: synthetic minority oversampling technique; XGB: extreme gradient boosting.

#### Table S6. Sensitivity analyses of the main comparison (all features, S1): tuning criterion, class weighting, and chain inputs

##### A. Hyperparameters tuned using AUC instead of the weighted F1-score

| | Weighted-F1 tuning (main analysis) | | | AUC tuning (sensitivity analysis) | | | | |
|---|---|---|---|---|---|---|---|---|
| **Algorithm** | **Unified** | **Single-label** | **Difference** | **Unified** | **Single-label** | **Difference** | **95% CI** | ***P*** |
| LR | 0.604 | 0.565 | +0.040 | 0.593 | 0.598 | −0.005 | −0.044 to 0.035 | .83 |
| RF | 0.643 | 0.648 | −0.004 | 0.650 | 0.652 | −0.002 | −0.018 to 0.014 | .88 |
| ET | 0.645 | 0.625 | +0.020 | 0.653 | 0.635 | +0.017 | −0.002 to 0.037 | .07 |
| XGB | 0.645 | 0.628 | +0.018 | 0.640 | 0.633 | +0.007 | −0.019 to 0.032 | .56 |
| GB | 0.635 | 0.627 | +0.008 | 0.628 | 0.617 | +0.011 | −0.017 to 0.039 | .42 |

##### B. Without class weights: unified vs single-label models

| Algorithm | Unified | Single-label | Difference | 95% CI | *P* |
|---|---|---|---|---|---|
| LR | 0.593 | 0.580 | +0.013 | −0.024 to 0.047 | .54 |
| RF | 0.642 | 0.639 | +0.003 | −0.010 to 0.017 | .61 |
| ET | 0.632 | 0.621 | +0.011 | −0.013 to 0.032 | .34 |
| XGB | 0.639 | 0.643 | −0.005 | −0.024 to 0.015 | .56 |
| GB | 0.610 | 0.621 | −0.011 | −0.037 to 0.015 | .37 |

##### C. Unified models passing predicted probabilities instead of predicted labels along the chain

| Algorithm | Probabilities | Predicted labels (main) | Difference | 95% CI | *P* |
|---|---|---|---|---|---|
| LR | 0.603 | 0.604 | −0.001 | −0.003 to 0.001 | .15 |
| RF | 0.643 | 0.643 | 0.000 | 0.000 to 0.000 | >.99 |
| ET | 0.618 | 0.645 | −0.027 | −0.054 to 0.000 | .052 |
| XGB | 0.644 | 0.645 | −0.002 | −0.009 to 0.007 | .66 |
| GB | 0.635 | 0.635 | 0.000 | 0.000 to 0.000 | >.99 |

Mean AUC across MCI, frailty, and fall risk; pooled out-of-fold predictions of the nested cross-validation (79 participants, 3 repeats; the same splits in all analyses). Difference: unified − single-label (A and B) or probabilities − predicted labels (C); 95% CIs and P values from the paired bootstrap (1000 resamples). A: for weighted-F1 tuning, the corresponding CIs and P values are given in Table S4 (stage S1). B: models identical to the main analysis except that class weights were removed; the class-weighted results are given in Table S4 (stage S1). C: unified models with class weights; only the information passed to the later steps of the chain was changed.
AUC: area under the receiver operating characteristic curve; ET: extra trees; GB: gradient boosting; LR: logistic regression; MCI: mild cognitive impairment; PR-AUC: area under the precision-recall curve; RF: random forest; SMOTE: synthetic minority oversampling technique; XGB: extreme gradient boosting.

#### Table S7. Performance of the unified and single-label approaches with nested model selection

| Metric | Outcome | Unified | Single-label | Difference (95% CI) | *P* |
|---|---|---|---|---|---|
| AUC | MCI | 0.741 | 0.751 | −0.010 (−0.064 to 0.044) | .76 |

| Metric | Outcome | Unified | Single-label | Difference (95% CI) | *P* |
|---|---|---|---|---|---|
| | Frailty | 0.568 | 0.517 | +0.051 (−0.017 to 0.116) | .13 |
| | Fall risk | 0.537 | 0.546 | −0.009 (−0.090 to 0.067) | .85 |
| | Mean | 0.615 | 0.605 | +0.011 (−0.034 to 0.051) | .58 |
| Weighted F1 | MCI | 0.686 | 0.692 | −0.006 (−0.060 to 0.044) | .86 |
| | Frailty | 0.649 | 0.636 | +0.013 (−0.030 to 0.055) | .60 |
| | Fall risk | 0.530 | 0.537 | −0.007 (−0.077 to 0.064) | .90 |
| | Mean | 0.622 | 0.622 | 0.000 (−0.035 to 0.034) | .95 |

In each outer training set, the combination of algorithm and stage (S1–S4) with the highest inner cross-validation weighted F1-score was selected: 1 combination for the unified approach and 1 per outcome for the single-label approach. Difference: unified − single-label; 95% CIs and P values from the paired bootstrap (1000 resamples).
AUC: area under the receiver operating characteristic curve; ET: extra trees; GB: gradient boosting; LR: logistic regression; MCI: mild cognitive impairment; PR-AUC: area under the precision-recall curve; RF: random forest; SMOTE: synthetic minority oversampling technique; XGB: extreme gradient boosting.

**Table S8.** Calibration of the final unified model

| Outcome | Observed prevalence | Mean predicted probability | Brier score | Reference Brier score | Calibration intercept | Calibration slope |
|---|---|---|---|---|---|---|
| MCI | 0.443 | 0.475 | 0.203 | 0.247 | −0.087 | 1.779 |
| Frailty | 0.278 | 0.422 | 0.220 | 0.201 | −0.780 | 0.589 |
| Fall risk | 0.418 | 0.478 | 0.248 | 0.243 | −0.294 | 0.382 |

Pooled out-of-fold predictions of the nested cross-validation (79 participants); values averaged over the 3 repeats. Reference Brier score=prevalence × (1 – prevalence). Ideal calibration: intercept 0 and slope 1.
AUC: area under the receiver operating characteristic curve; MCI: mild cognitive impairment; NPV: negative predictive value; PPV: positive predictive value; PR-AUC: area under the precision-recall curve.

Table S9. Performance of the final unified model for each outcome, with 95% CIs

| Outcome | Prevalence | AUC | PR-AUC | Sensitivity | Specificity | PPV | NPV | Weighted F1 |
|---|---|---|---|---|---|---|---|---|
| MCI | 0.443 | 0.779 (0.680–0.872) | 0.684 (0.527–0.855) | 0.648 (0.491–0.784) | 0.818 (0.703–0.927) | 0.740 (0.566–0.887) | 0.745 (0.624–0.847) | 0.740 (0.646–0.830) |
| Frailty | 0.278 | 0.606 (0.456–0.750) | 0.414 (0.260–0.624) | 0.500 (0.314–0.706) | 0.789 (0.686–0.879) | 0.478 (0.270–0.670) | 0.804 (0.704–0.899) | 0.711 (0.618–0.800) |
| Fall risk | 0.418 | 0.551 (0.444– | 0.490 (0.360– | 0.414 (0.282– | 0.674 (0.566– | 0.478 (0.313– | 0.616 (0.495– | 0.560 (0.474– |

| Outcome | Prevalence | AUC | PR-AUC | Sensitivity | Specificity | PPV | NPV | Weighted F1 |
|---|---|---|---|---|---|---|---|---|
| | | 0.653) | 0.641) | 0.552) | 0.783) | 0.631) | 0.747) | 0.647) |
| **Mean** | – | 0.645 (0.569–0.722) | 0.529 (0.433–0.663) | 0.521 (0.398–0.639) | 0.761 (0.681–0.838) | 0.565 (0.449–0.676) | 0.721 (0.651–0.788) | 0.670 (0.614–0.721) |

Pooled out-of-fold predictions of the nested cross-validation (79 participants, 3 repeats); 95% CIs from the bootstrap (1000 resamples). All metrics except AUC and PR-AUC at a threshold of 0.5. The prevalence of each outcome is the chance-level PR-AUC.

AUC: area under the receiver operating characteristic curve; MCI: mild cognitive impairment; NPV: negative predictive value; PPV: positive predictive value; PR-AUC: area under the precision-recall curve.

**Figure S2.** Performance of the final unified model for each outcome with 95% bootstrap CIs

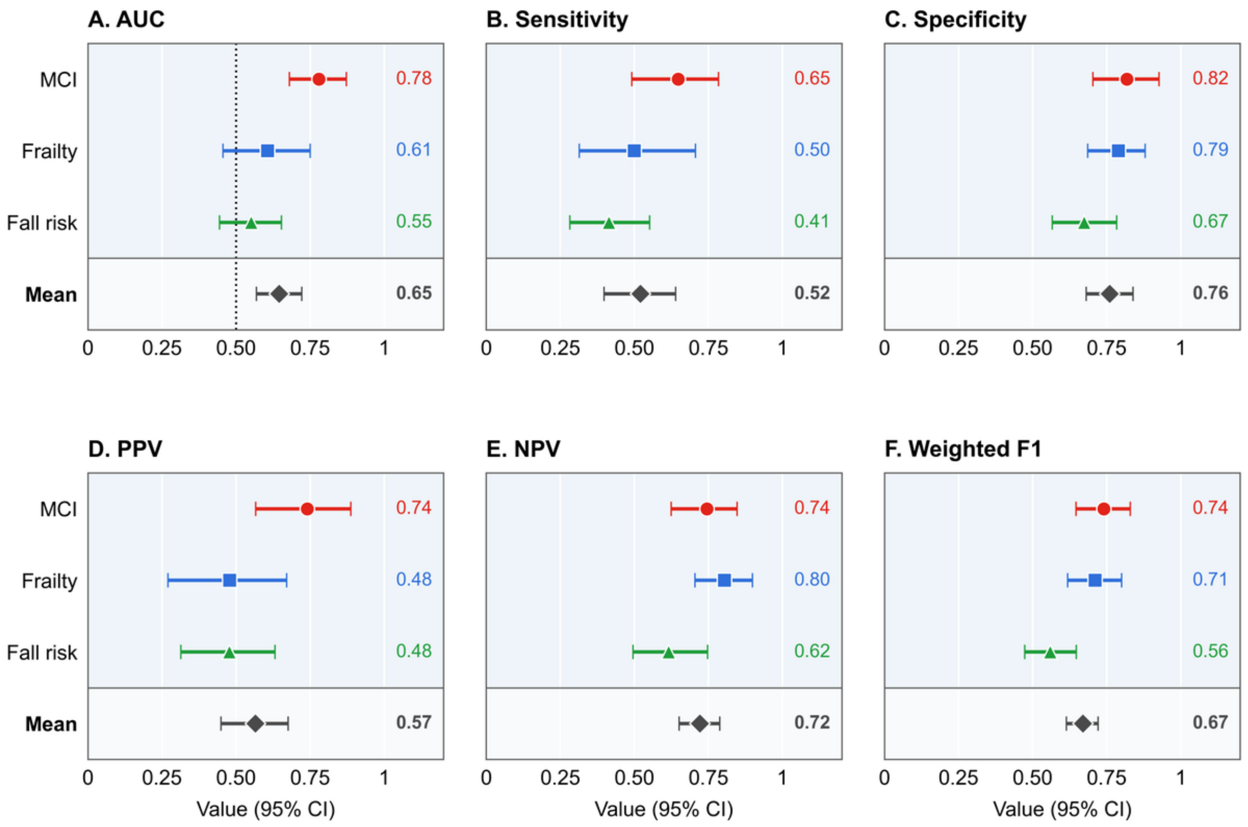


The dashed line marks chance-level AUC (0.5). AUC: area under the receiver operating characteristic curve; MCI: mild cognitive impairment; PR-AUC: area under the precision-recall curve.

**Figure S3.** Confusion matrices of the final unified model at a threshold of 0.5 (n=79)

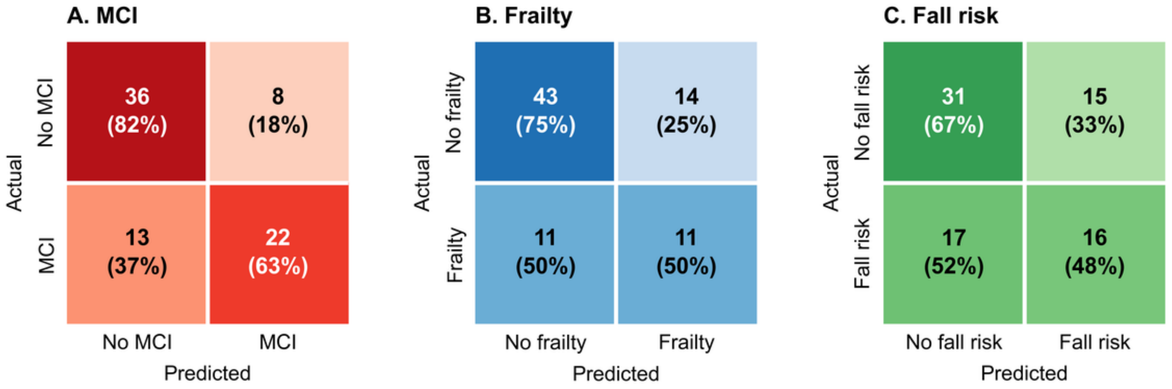


Each participant's predicted probability was averaged over the 3 repeats and classified at a threshold of 0.5; cells show the number of participants and the percentage of each actual group. Because the probabilities were averaged before classification, sensitivity and specificity can differ slightly from Table 5, where the metrics were averaged over the repeats. MCI: mild cognitive impairment.

#### Multimedia Appendix. Supplementary interpretation and robustness results

**Figure S4.** SHAP analysis of the final unified model in the training participants. (A) The 20 most important features. Each dot is 1 participant; dots to the right of 0 indicate a higher predicted risk. Color shows the feature value, from low (blue) to high (pink); for binary features, pink indicates female or yes. (B) Share of each outcome in each feature's importance, with the feature's share of total importance at the right.

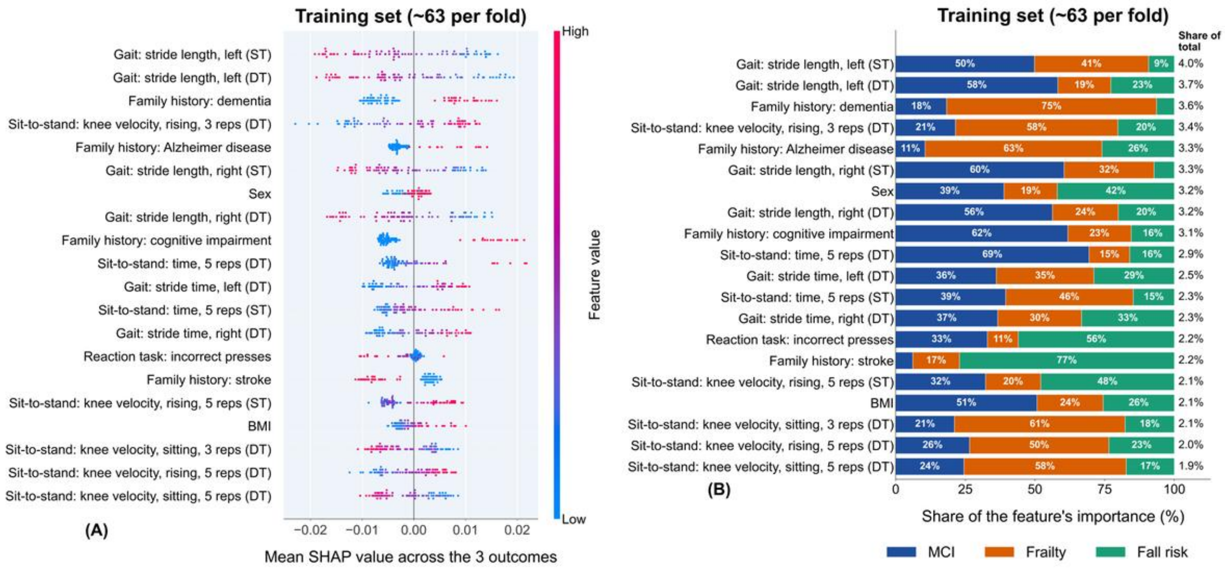


#### Table S10. SHAP importance of all 59 features and the share of each outcome in the final unified model

| Rank (training) | Rank (test) | Feature | Share of total, % | MCI, % | Frailty, % | Fall risk, % | Pattern |
|---|---|---|---|---|---|---|---|
| 1 | 1 | Gait: stride length, left (ST) | 4.0 | 49.8 | 41.0 | 9.2 | Shared |
| 2 | 3 | Gait: stride length, left (DT) | 3.7 | 58.2 | 19.0 | 22.8 | Shared |
| 3 | 2 | Family history: dementia | 3.6 | 18.2 | 75.4 | 6.4 | Single (Frailty) |
| 4 | 6 | Sit-to-stand: knee velocity, rising, 3 reps (DT) | 3.4 | 21.4 | 58.3 | 20.3 | Shared |
| 5 | 4 | Family history: Alzheimer disease | 3.3 | 10.6 | 63.5 | 26.0 | Single (Frailty) |
| 6 | 7 | Gait: stride length, right (ST) | 3.3 | 60.4 | 32.3 | 7.3 | Single (MCI) |
| 7 | 5 | Sex | 3.2 | 38.9 | 19.1 | 42.0 | Shared |
| 8 | 8 | Gait: stride length, right (DT) | 3.2 | 56.3 | 23.6 | 20.1 | Shared |
| 9 | 9 | Family history: cognitive impairment | 3.1 | 61.8 | 22.7 | 15.5 | Single (MCI) |

| Rank (training) | Rank (test) | Feature | Share of total, % | MCI, % | Frailty, % | Fall risk, % | Pattern |
|---|---|---|---|---|---|---|---|
| 10 | 10 | Sit-to-stand: time, 5 reps (DT) | 2.9 | 69.4 | 14.6 | 16.0 | Single (MCI) |
| 11 | 11 | Gait: stride time, left (DT) | 2.5 | 36.1 | 35.0 | 28.9 | Shared |
| 12 | 14 | Sit-to-stand: time, 5 reps (ST) | 2.3 | 39.5 | 45.8 | 14.7 | Shared |
| 13 | 12 | Gait: stride time, right (DT) | 2.3 | 36.6 | 30.1 | 33.3 | Shared |
| 14 | 15 | Reaction task: incorrect presses | 2.2 | 32.9 | 11.1 | 56.0 | Shared |
| 15 | 13 | Family history: stroke | 2.2 | 6.1 | 16.8 | 77.1 | Single (Fall risk) |
| 16 | 18 | Sit-to-stand: knee velocity, rising, 5 reps (ST) | 2.1 | 32.2 | 19.8 | 47.9 | Shared |
| 17 | 16 | BMI | 2.1 | 50.7 | 23.7 | 25.6 | Shared |
| 18 | 19 | Sit-to-stand: knee velocity, sitting, 3 reps (DT) | 2.1 | 21.0 | 61.3 | 17.7 | Single (Frailty) |
| 19 | 20 | Sit-to-stand: knee velocity, rising, 5 reps (DT) | 2.0 | 26.5 | 50.0 | 23.5 | Shared |
| 20 | 21 | Sit-to-stand: knee velocity, sitting, 5 reps (DT) | 1.9 | 24.5 | 58.2 | 17.3 | Shared |
| 21 | 17 | Family history: alcoholism | 1.9 | 6.8 | 22.2 | 71.0 | Single (Fall risk) |
| 22 | 26 | Age | 1.9 | 47.6 | 18.6 | 33.8 | Shared |
| 23 | 25 | Balance: CoP mean velocity, AP (ST) | 1.9 | 59.4 | 20.8 | 19.8 | Shared |
| 24 | 24 | Serial subtraction: total responses, 15 s (DT) | 1.8 | 17.0 | 22.1 | 60.9 | Single (Fall risk) |
| 25 | 22 | Balance: AUC, AP (ST) | 1.8 | 38.0 | 39.0 | 23.0 | Shared |
| 26 | 30 | Sit-to-stand: time, 3 reps (ST) | 1.8 | 60.4 | 28.9 | 10.6 | Single (MCI) |
| 27 | 23 | Weight | 1.8 | 39.1 | 11.1 | 49.7 | Shared |
| 28 | 27 | Family history: bipolar disorder | 1.8 | 60.1 | 36.9 | 3.0 | Single (MCI) |
| 29 | 32 | Sit-to-stand: knee velocity, rising, 3 reps (ST) | 1.7 | 36.5 | 31.5 | 32.1 | Shared |
| 30 | 31 | Reaction task: reaction time | 1.6 | 63.8 | 25.4 | 10.7 | Single (MCI) |
| 31 | 29 | Gait: stride time, left (ST) | 1.6 | 35.9 | 28.6 | 35.4 | Shared |
| 32 | 33 | Sit-to-stand: knee velocity, sitting, 5 reps (ST) | 1.6 | 19.8 | 41.6 | 38.6 | Shared |

| Rank (training) | Rank (test) | Feature | Share of total, % | MCI, % | Frailty, % | Fall risk, % | Pattern |
|---|---|---|---|---|---|---|---|
| 33 | 28 | Reaction task: buttons pressed | 1.6 | 62.6 | 28.3 | 9.1 | Single (MCI) |
| 34 | 34 | Height | 1.5 | 20.1 | 30.6 | 49.2 | Shared |
| 35 | 38 | Sit-to-stand: time, 3 reps (DT) | 1.3 | 61.5 | 29.1 | 9.4 | Single (MCI) |
| 36 | 35 | Serial subtraction: incorrect responses, 15 s (DT) | 1.3 | 70.5 | 15.9 | 13.6 | Single (MCI) |
| 37 | 36 | Balance: CoP 95% sway area (DT) | 1.2 | 11.5 | 71.4 | 17.1 | Single (Frailty) |
| 38 | 37 | Balance: CoP mean velocity (ST) | 1.2 | 47.5 | 27.0 | 25.5 | Shared |
| 39 | 40 | Sit-to-stand: knee velocity, sitting, 3 reps (ST) | 1.2 | 39.0 | 35.0 | 26.1 | Shared |
| 40 | 39 | Gait: stride time, right (ST) | 1.2 | 22.4 | 42.8 | 34.9 | Shared |
| 41 | 42 | Balance: CoP mean velocity, AP (DT) | 1.1 | 20.6 | 48.4 | 31.0 | Shared |
| 42 | 41 | Balance: AUC, AP (DT) | 1.1 | 43.8 | 19.7 | 36.5 | Shared |
| 43 | 45 | Balance: AUC, ML (DT) | 1.0 | 27.2 | 38.3 | 34.5 | Shared |
| 44 | 43 | Step-down: CoM ML displacement, right | 0.9 | 26.1 | 43.3 | 30.6 | Shared |
| 45 | 46 | Step-down: trail time, right | 0.8 | 57.3 | 15.9 | 26.7 | Shared |
| 46 | 44 | Step-down: trail time, left | 0.8 | 39.9 | 37.6 | 22.5 | Shared |
| 47 | 47 | Balance: CoP 95% sway area (ST) | 0.8 | 29.2 | 38.9 | 32.0 | Shared |
| 48 | 49 | Balance: CoP mean velocity (DT) | 0.8 | 22.9 | 39.5 | 37.6 | Shared |
| 49 | 48 | Step-down: CoM ML displacement, left | 0.8 | 20.0 | 35.6 | 44.5 | Shared |
| 50 | 50 | Balance: AUC, ML (ST) | 0.7 | 32.3 | 43.5 | 24.2 | Shared |
| 51 | 51 | Balance: CoP mean velocity, ML (ST) | 0.7 | 20.9 | 22.3 | 56.8 | Shared |
| 52 | 52 | Serial subtraction: interference, total (DT) | 0.6 | 39.3 | 25.1 | 35.7 | Shared |
| 53 | 53 | Step-down: lead time, left | 0.6 | 25.4 | 49.8 | 24.9 | Shared |
| 54 | 54 | Step-down: lead time, right | 0.5 | 41.3 | 32.5 | 26.1 | Shared |
| 55 | 55 | Balance: CoP mean velocity, ML (DT) | 0.5 | 22.3 | 31.9 | 45.8 | Shared |
| 56 | 56 | Family history: schizophrenia | 0.4 | 56.2 | 8.6 | 35.2 | Shared |
| 57 | 58 | Serial subtraction: interference, correct (DT) | 0.4 | 27.5 | 37.6 | 34.9 | Shared |

| Rank (training) | Rank (test) | Feature | Share of total, % | MCI, % | Frailty, % | Fall risk, % | Pattern |
|---|---|---|---|---|---|---|---|
| 58 | 57 | Family history: Parkinson disease | 0.4 | 32.2 | 36.4 | 31.5 | Shared |
| 59 | 59 | Race | 0.1 | 16.8 | 53.8 | 29.4 | Shared |

Final unified model (extra trees, all features); SHAP values from nested cross-validation, averaged over the 3 repeats (training set: ~63 participants per fold; test set: ~16 participants per fold). Features are listed by their total importance in the training set; Rank (test) is the rank of the same feature in the test set. Share of total: the feature's mean absolute SHAP value summed over the 3 outcomes, as a percentage of the sum over all 59 features. MCI, frailty, and fall risk: each outcome's share of the feature's importance in the training set (rows sum to 100%). All features contributed to all 3 outcomes. Single (outcome): 1 outcome accounted for 60% or more of the feature's importance; shared: otherwise.

AP: anteroposterior; AUC: area under the curve; BMI: body mass index; CoM: center of mass; CoP: center of pressure; DT: dual task; MCI: mild cognitive impairment; ML: mediolateral; reps: repetitions; SHAP: Shapley additive explanations; ST: single task.

#### Table S11. Summary of effect sizes of the 59 features between participants with and without each condition

| Outcome | With / without | Median \|SMD\| | ≥ 0.2 | ≥ 0.5 | ≥ 0.8 | Largest effect (\|SMD\|, direction with the condition) |
|---|---|---|---|---|---|---|
| MCI | 35 / 44 | 0.383 | 42 | 22 | 5 | Gait: stride length, left (ST) (1.08, shorter) |
| Frailty | 22 / 57 | 0.349 | 43 | 14 | 1 | Sit-to-stand: knee velocity, rising, 3 reps (DT) (0.90, slower rising) |
| Fall risk | 33 / 46 | 0.156 | 21 | 0 | 0 | Gait: stride length, left (DT) (0.41, shorter) |

All participants (n=79); missing values imputed with the median. SMD: Hedges g for 49 continuous features and SMD for proportions for 10 binary features. Columns ≥0.2, ≥0.5, and ≥0.8 give the number of features with at least a small, medium, or large effect, respectively. Effect sizes for every feature are given in Table S12.
DT: dual task; MCI: mild cognitive impairment; reps: repetitions; SMD: standardized mean difference; ST: single task.

#### Table S12. Effect sizes (SMD) of all 59 features between participants with and without each condition

| Feature | MCI | Frailty | Fall risk |
|---|---|---|---|
| *Demographics* | | | |
| Age | +0.45 | −0.01 | −0.34 |

| Feature | MCI | Frailty | Fall risk |
|---|---|---|---|
| Height | −0.07 | −0.37 | −0.20 |
| Weight | **+0.54** | +0.14 | −0.34 |
| BMI | **+0.65** | +0.40 | −0.31 |
| Sex† | −0.35 | +0.21 | +0.32 |
| Race† | +0.04 | +0.16 | +0.05 |
| *Family history* | | | |
| Alzheimer disease† | +0.24 | +0.48 | −0.26 |
| Dementia† | +0.22 | +0.46 | −0.03 |
| Parkinson disease† | +0.02 | −0.01 | +0.06 |
| Stroke† | +0.06 | +0.03 | −0.30 |
| Cognitive impairment† | +0.44 | +0.34 | +0.11 |
| Schizophrenia† | −0.38 | +0.05 | +0.20 |
| Bipolar disorder† | **+0.52** | −0.23 | +0.01 |
| Alcoholism† | −0.02 | +0.22 | −0.19 |
| *Gait* | | | |
| Stride length, left (ST) | **−1.08** | **−0.71** | −0.13 |
| Stride length, right (ST) | **−1.02** | **−0.59** | −0.02 |
| Stride time, left (ST) | +0.37 | +0.35 | +0.08 |
| Stride time, right (ST) | +0.34 | +0.39 | +0.08 |
| Stride length, left (DT) | **−1.06** | **−0.55** | −0.41 |
| Stride length, right (DT) | **−1.02** | **−0.56** | −0.38 |
| Stride time, left (DT) | **+0.62** | **+0.54** | +0.29 |
| Stride time, right (DT) | **+0.62** | +0.48 | +0.26 |
| *Sit-to-stand* | | | |
| Time, 3 reps (ST) | **+0.77** | **+0.64** | +0.16 |
| Knee velocity, rising, 3 reps (ST) | **+0.60** | +0.50 | +0.32 |
| Knee velocity, sitting, 3 reps (ST) | **−0.60** | **−0.55** | −0.20 |
| Time, 5 reps (ST) | **+0.78** | **+0.64** | +0.24 |
| Knee velocity, rising, 5 reps (ST) | **+0.63** | **+0.51** | +0.37 |
| Knee velocity, sitting, 5 reps (ST) | **−0.59** | **−0.51** | −0.31 |
| Time, 3 reps (DT) | **+0.72** | +0.35 | −0.05 |
| Knee velocity, rising, 3 reps (DT) | **+0.59** | **+0.90** | +0.37 |
| Knee velocity, sitting, 3 reps (DT) | −0.44 | **−0.74** | −0.34 |
| Time, 5 reps (DT) | **+0.95** | +0.50 | −0.10 |
| Knee velocity, rising, 5 reps (DT) | **+0.62** | **+0.68** | +0.31 |
| Knee velocity, sitting, 5 reps (DT) | −0.47 | **−0.64** | −0.32 |
| *Static balance* | | | |
| CoP 95% sway area (ST) | +0.19 | +0.20 | +0.17 |
| CoP mean velocity (ST) | +0.42 | +0.26 | +0.15 |
| CoP mean velocity, ML (ST) | +0.17 | +0.04 | +0.13 |
| CoP mean velocity, AP (ST) | +0.49 | +0.32 | +0.15 |
| AUC, AP (ST) | **+0.52** | +0.29 | +0.12 |
| AUC, ML (ST) | +0.13 | −0.05 | +0.03 |
| CoP 95% sway area (DT) | +0.20 | +0.35 | +0.01 |
| CoP mean velocity (DT) | +0.27 | +0.25 | +0.16 |
| CoP mean velocity, ML (DT) | +0.03 | −0.02 | +0.06 |
| CoP mean velocity, AP (DT) | +0.31 | +0.30 | +0.17 |

| Feature | MCI | Frailty | Fall risk |
|---|---|---|---|
| AUC, AP (DT) | +0.41 | +0.21 | +0.01 |
| AUC, ML (DT) | +0.00 | −0.03 | +0.04 |
| *Step-down* | | | |
| Lead time, left | +0.17 | −0.06 | −0.08 |
| Trail time, left | +0.20 | +0.20 | +0.00 |
| CoM ML displacement, left | −0.02 | −0.04 | −0.10 |
| Lead time, right | +0.15 | +0.14 | +0.19 |
| Trail time, right | +0.38 | +0.32 | +0.30 |
| CoM ML displacement, right | +0.01 | −0.06 | −0.14 |
| *Reaction time* | | | |
| Reaction time | **+0.74** | +0.45 | +0.14 |
| Incorrect presses | +0.07 | +0.39 | −0.02 |
| Buttons pressed | **−0.73** | −0.48 | −0.14 |
| *Serial subtraction (dual task)* | | | |
| Total responses, 15 s (DT) | −0.21 | −0.46 | −0.36 |
| Incorrect responses, 15 s (DT) | +0.30 | −0.09 | +0.03 |
| Interference, total (DT) | −0.02 | −0.05 | +0.12 |
| Interference, correct (DT) | −0.20 | +0.40 | −0.19 |

All participants (n=79; MCI 35 vs 44, frailty 22 vs 57, fall risk 33 vs 46); missing values imputed with the median. Values are SMDs (Hedges g for continuous features; SMD for proportions for binary features, marked †). Positive values indicate a higher value or proportion among participants with the condition. Knee velocity while rising is negative in this dataset, so a positive value indicates slower rising. Bold indicates at least a medium effect (|SMD|≥0.5). Binary coding: sex, 0=male, 1=female; race, 0=White, 1=non-White; family history, 0=no, 1=yes. DT: dual task; MCI: mild cognitive impairment; reps: repetitions; SMD: standardized mean difference; ST: single task. AP: anteroposterior; BMI: body mass index; CoM: center of mass; CoP: center of pressure; ML: mediolateral.

#### Table S13. Chain-input importance of the final unified model on the test data

| Chain step and input | Decrease in test AUC, mean (SD) | Rank among all inputs of the step | Most important original feature of the step (decrease in test AUC) |
|---|---|---|---|
| Frailty step: predicted MCI | 0.006 (0.008) | 5 of 60 | Sit-to-stand: knee velocity, rising, 3 reps (DT) (0.011) |
| Fall-risk step: predicted MCI | 0.004 (0.005) | 17 of 61 | Reaction task: incorrect presses (0.017) |
| Fall-risk step: predicted frailty | 0.062 (0.013) | 1 of 61 | Reaction task: incorrect presses (0.017) |

Final unified model (extra trees, all features, S1); pooled out-of-fold test predictions of the nested cross-validation (79 participants, 3 repeats). In each of the 15 outer folds, each input of the frailty and fall-risk steps was shuffled 20 times among the test participants, and the decrease in test AUC was averaged over the 3 repeats (SD across shuffles). The frailty step had 60 inputs (59 features and predicted MCI), and the fall-risk step had 61 (59 features, predicted MCI, and predicted frailty).
AUC: area under the receiver operating characteristic curve; DT: dual task; MCI: mild cognitive impairment; reps: repetitions.

#### Table S14. Test AUC of the final unified model after removing 1 feature domain at a time

| | Test AUC | P (vs final model) |
|---|---|---|

| Configuration | Features | MCI | Frailty | Fall risk | Mean | Change in mean AUC (95% CI) | MCI | Frailty | Fall risk | Mean |
|---|---|---|---|---|---|---|---|---|---|---|
| Final model (all features) | 59 | 0.779 | 0.606 | 0.551 | 0.645 | — | — | — | — | — |
| Without demographics | 53 | 0.774 | 0.620 | 0.522 | 0.638 | −0.007 (−0.034 to 0.021) | .64 | .51 | .34 | .63 |
| Without family history | 51 | 0.772 | 0.623 | 0.573 | 0.656 | +0.010 (−0.015 to 0.038) | .66 | .57 | .47 | .43 |
| Without gait | 51 | 0.749 | 0.587 | 0.536 | 0.624 | −0.022 (−0.058 to 0.013) | .26 | .60 | .65 | .24 |
| Without reaction time | 56 | 0.772 | 0.618 | 0.560 | 0.650 | +0.004 (−0.016 to 0.026) | .54 | .53 | .68 | .68 |
| Without sit-to-stand | 47 | 0.781 | 0.638 | 0.547 | 0.655 | +0.010 (−0.025 to 0.045) | .87 | .39 | .91 | .59 |
| Without static balance | 47 | 0.794 | 0.607 | 0.563 | 0.655 | +0.009 (−0.020 to 0.039) | .32 | .99 | .69 | .53 |
| Without serial subtraction (dual task) | 55 | 0.781 | 0.632 | 0.574 | 0.662 | +0.017 (−0.002 to 0.039) | .85 | .11 | .35 | .08 |
| Without step-down | 53 | 0.780 | 0.620 | 0.574 | 0.658 | +0.012 (−0.008 to 0.033) | .96 | .40 | .28 | .25 |
| Sensor features only | 45 | 0.766 | 0.639 | 0.529 | 0.645 | −0.001 (−0.031 to 0.029) | .51 | .28 | .56 | .96 |

Extra trees with all remaining features (S1); pooled out-of-fold test predictions of the same nested cross-validation as the final model (79 participants; values averaged over the 3 repeats). Change in mean AUC=model without the domain − final model. 95% CIs and P values from the paired bootstrap (1000 resamples). Motor and cognitive-task features only: demographic and family-history variables left out.
AUC: area under the receiver operating characteristic curve; MCI: mild cognitive impairment.

#### Table S15. Test performance of the unified model for each order of the outcomes in the classifier chain, with comparisons against the final order

| Chain order | Test AUC | | | | Mean weighted F1 | P (vs final order) | | | |
|---|---|---|---|---|---|---|---|---|---|
| | MCI | Frailty | Fall risk | Mean | | MCI | Frailty | Fall risk | Mean |
| MCI → frailty → fall risk (final model) | 0.779 | 0.606 | 0.551 | 0.645 | 0.670 | — | — | — | — |
| MCI → fall risk → frailty | 0.777 | 0.593 | 0.505 | 0.625 | 0.644 | .82 | .50 | .15 | .13 |
| Frailty → MCI → fall risk | 0.785 | 0.602 | 0.539 | 0.642 | 0.656 | .56 | .78 | .55 | .65 |
| Frailty → fall risk → MCI | 0.780 | 0.611 | 0.560 | 0.650 | 0.677 | .94 | .77 | .62 | .63 |
| Fall risk → MCI → frailty | 0.776 | 0.618 | 0.505 | 0.633 | 0.653 | .79 | .50 | .11 | .29 |
| Fall risk → frailty → MCI | 0.785 | 0.634 | 0.493 | 0.637 | 0.646 | .57 | .17 | .03[a] | .54 |

Extra trees with all features (S1); pooled out-of-fold test predictions of the same nested cross-validation for every order (79 participants; values averaged over the 3 repeats). *P* values from the paired bootstrap (1000 resamples) comparing each order with the final order.
[a]*P*<.05 compared with the final order.
AUC: area under the receiver operating characteristic curve; MCI: mild cognitive impairment.

### Multimedia Appendix. Supplementary sample-size results

#### Table S16. Sensitivity analysis: detectable effect sizes, power, and required sample sizes

| Outcome | With / without | Detectable \|SMD\| (80% power) | Power | | | Total sample size needed (80% power) | | |
|---|---|---|---|---|---|---|---|---|
| | | | Small (0.2) | Medium (0.5) | Large (0.8) | Small (0.2) | Medium (0.5) | Large (0.8) |
| MCI | 35 / 44 | 0.64 | 0.14 | 0.59 | 0.94 | 798 | 130 | 52 |
| Frailty | 22 / 57 | 0.71 | 0.12 | 0.50 | 0.88 | 979 | 159 | 64 |
| Fall risk | 33 / 46 | 0.65 | 0.14 | 0.58 | 0.93 | 809 | 132 | 53 |

Power for a difference between 2 independent groups (two-sided, α=.05) with the group sizes of the 79 participants. Detectable |SMD| is the smallest absolute standardized mean difference detectable with 80% power. Required sample sizes are totals and keep the observed proportion of participants with each condition. Small, medium, and large effects correspond to |SMD| of 0.2, 0.5, and 0.8.

MCI: mild cognitive impairment; SMD: standardized mean difference.
